# Adjusting for the progressive digitization of health records: working examples on a multi-hospital clinical data warehouse

**DOI:** 10.1101/2023.08.17.23294220

**Authors:** Adam Remaki, Benoît Playe, Paul Bernard, Simon Vittoz, Matthieu Doutreligne, Gilles Chatelier, Etienne Audureau, Emmanuelle Kempf, Raphaël Porcher, Romain Bey

## Abstract

**Objectives:** To propose a new method to account for time-dependent data missingness caused by the increasing digitization of health records in the analysis of large-scale clinical data.

**Materials and Methods:** Following a data-driven approach we modeled the progressive adoption of a common electronic health record in 38 hospitals. To this end, we analyzed data collected between 2013 and 2022 and made available in the clinical data warehouse of the Greater Paris University Hospitals. Depending on the category of data, we worked either at the hospital, department or unit level. We evaluated the performance of this model with a retrospective cohort study. We measured the temporal variations of some quality and epidemiological indicators by successively applying two methods, either a naive analysis or a novel complete-source-only analysis that accounts for digitization-induced missingness.

**Results:** Unrealistic temporal variations of quality and epidemiological indicators were observed when a naive analysis was performed, but this effect was either greatly reduced or disappeared when the complete-source-only method was applied.

**Discussion:** We demonstrated that a data-driven approach can be used to account for missingness induced by the progressive digitization of health records. This work focused on hospitalization, emergency department and intensive care units records, along with diagnostic codes, discharge prescriptions and consultation reports. Other data categories may require specific modeling of their associated data sources.

**Conclusions:** Electronic health records are constantly evolving and new methods should be developed to debias studies that use these unstable data sources.

## 1 Background and Significance

The ongoing digitization of healthcare is generating more and more routine data. These data contain rich and diverse information about patients, opening up new prospects for research, innovation, quality monitoring and public health surveillance.[1–4] However, their analysis also raises new methodological challenges that differ in many ways from those associated with data collected specifically for research purposes.[5–8] In particular, the rapid pace of digitization leads to frequent variations in data availability, format, and coverage, thereby hampering the conduct of multicenter longitudinal studies, as records collected in different contexts are not always comparable.[9–22] To address this pervasive problem, some studies have proposed tools to facilitate the detection of drifts or sudden changes in statistical distributions,[9, 23–26] while others have explored methods to homogenize datasets before their analysis, such as imputing missing data or dropping incomplete cases.[27–29] These seminal studies have paved the way for the analysis of routine data, but many issues remain to be addressed.

First, temporal variations in statistical distributions can be induced by technological changes, but also by other mechanisms. Disentangling technology-induced drifts from variations caused, for example, by reorganizations of care or changes in patients populations seems to be a prerequisite for answering many questions of interest.

Second, the technological changes that need to be accounted for are increasingly complex. For example, clinical data warehouses (CDWs) include more and more healthcare sites, and within those sites, each department may have its own characteristics. In addition, the data being analyzed are increasingly diverse, including diagnostic codes, clinical reports, laboratory tests, imaging, medications, etc., thus multiplying the number of dynamics to consider. Controlling these complex drifts of data distributions by mere visual inspection would therefore mobilize enormous resources, and there is a clear need for some degree of automation.[29]

## 2 Objective

In this study, we developed and evaluated a new method to automatically handle time-dependent missingness induced by the progressive digitization of hospital health records. We analyzed data collected in the Greater Paris University Hospitals (Assistance Publique-Hôpitaux de Paris, APHP). Among the many mechanisms that may induce missing data, we focused on the effect of the progressive adoption of functionalities used to collect administrative records, diagnostic codes, discharge prescriptions, and consultation reports. Using a data-driven approach, we modeled the pace of their adoption at different levels (hospitals, departments, units). We then evaluated the performance of this detailed model by measuring some quality and epidemiological indicators. We evaluated their stability over time using two different methods, either a naive approach that did not take into account the progressive digitization of health records, or a novel approach, complete-source-only analysis, that took advantage of the previous modeling of this mechanism. This work aimed to answer the following questions:

- Can we automatically model the progressive digitization of health records using a data-only approach?
- Can we use such a model to adjust for the time-dependent missingness induced by this mechanism when conducting archetypal observational studies?

## 3 Materials and Methods

The study was reviewed and approved by the institutional review board of the AP-HP (IRB00011591, decision CSE21-33). French regulations do not require written patient consent for this type of research. In accordance with the European General Data Protection Regulation, patients were informed and those who objected to the secondary use of their data for research were excluded from the study. This report follows the REporting of studies Conducted using Observational Routinely-collected health Data (RECORD) reporting guideline in the Section F of the Supplementary Materials.[30]

### 3.1 Data source

AP-HP includes 38 hospitals spread across the Paris region (22 000 beds, 1.5 million hospitalizations per year). A common electronic health record (EHR) software, ORBIS Dedalus Healthcare, has been progressively adopted since 2012. Each of the six data categories considered in this study is collected using a different EHR functionality (i.e., administrative records related to hospitalizations, emergency departments -ED- or intensive care units -ICU-, diagnostic codes, discharge prescriptions, and consultation reports). The CDW follows the Observational Medical Outcomes Partnership-Common Data Model version 5.4 standard.[31] Data are integrated into the CDW on a daily basis, and this study was conducted on July 31^st^, 2023.

### 3.2 Modeling of the EHR adoption

The adoption of AP-HP’s common EHR is done by functionality, and the collection of each category of data depends on the use of a particular functionality. The functionalities dedicated to hospitalization records are adopted at the hospital level, while other functionalities are typically adopted at the department or unit level (e.g., ED or ICU visits, diagnostic codes, or clinical reports). No curated knowledge base currently provides information on the effective use of each EHR functionality, so we took a data-driven approach to generate it. Extracting the dynamics of EHR adoption from data is not straightforward because of the interplay of different mechanisms: i) the technical introduction of EHR functionalities causes a sudden increase in data availability, but ii) their gradual adoption by clinicians smooths this effect, which is also sometimes blurred by iii) the copying of data collected in an earlier software into the new EHR. Such mechanisms influence the shape of data availability curves, which rarely follow an ideal step shape when plotted over time. However, compared to variations induced by other mechanisms such as changes in clinical practice, the introduction and adoption of new EHR functionalities usually induces abrupt variations in data availability that are more localized in time and within healthcare sites. Therefore, to automatically detect EHR adoption we computed *c*(*t*), an estimate of completeness per EHR functionality and per healthcare site (i.e., hospital, department, or unit) for month *t*, and we fitted step functions to these abruptly changing time series (see Section A.1 of the Supplementary Materials for details).

We adopted two definitions of *c*(*t*) depending on the EHR functionality: either the proportion of hospitalization records with at least one data point (to study the EHR functionalities used to collect diagnostic codes and discharge prescriptions) or, when such a denominator was not available, the monthly number of data points divided by its highest value measured during the study period (to study hospitalization records, emergency records, intra-hospitalization visits to ICUs and consultation reports). This modeling provided, for each healthcare site and each EHR functionality, *t*_0_, the estimated date of adoption, and *c*_0_, the stabilized average value of completeness after that date. In the case of visits to ICUs, we also evaluated an alternative modeling that consisted of fitting rectangular functions to the completeness estimates instead of step functions, thus additionally providing *t*_1_, an extinction date corresponding to the end of data collection in a unit. This alternative modeling was motivated by the observation that data collected in a single ICU sometimes disappeared after a certain date due to hospital reorganization (see Figure S1 in the Supplement). Finally, to assess the goodness of fit of our models, we computed an *error* term defined by the mean squared error between *c*_0_ and *c*(*t*) after *t*_0_ (see Figure S2 in the Supplement).

### 3.3 Quality and epidemiological indicators

EHR data can be used to study retrospectively or to monitor prospectively quality or epidemiological indicators. However, time-dependent missingness can lead to biased estimates of these indicators. In this article, quality indicators were defined as the monthly proportion of hospitalizations for which some outcomes were observed, and epidemiological indicators were defined as the weekly number of hospitalizations related to some seasonal epidemics. Table 1 lists the indicators we considered and the categories of data we used to select each cohort and calculate the outcomes. Bronchiolotis-related and flu-related hospitalizations were selected using *J*21 and *J*09, *J*10, *J*11 International Classification of Diseases 10^th^ Revision (ICD-10) codes, respectively.

**Table 1:** Quality indicators and epidemiological indicators.

| Indicator | Definition | EHR functionality used for cohort selection (adoption level) | EHR functionalities used for outcome measurement (adoption level) |
| --- | --- | --- | --- |
| 30-day rehospitalization | Proportion of hospitalizations with a rehospitalization in the 30 days after discharge.[32, 33] | Hospitalization record (hospital-level) | Hospitalization record (hospital-level) |
| 30-day ED consultation* | Proportion of hospitalizations with at least one consultation in ED in the 30 days after discharge. | Hospitalization record (hospital-level) | Emergency record (hospital-level) |
| 30-day consultation | Proportion of hospitalizations with at least one outpatient visit occurring in the 30 days after discharge.[34] | Hospitalization record (hospital-level) | Consultation reports (department-level) |
| Discharge prescription | Proportion of hospitalizations with at least one discharge prescription delivered to the patient. | Hospitalization record (hospital-level) | Discharge prescription (department-level) |
| 30-day ICU readmission* | Proportion of hospitalizations with at least one readmission in an ICU in the 30 days after discharge.[35] | Hospitalization record (hospital-level) | Intra-hospitalization visit to ICU* (unit-level) |
| Bronchiolitis-related hospitalizations | Weekly number of hospitalizations with a bronchiolitis diagnosis.[36] | Hospitalization record (hospital-level) | Diagnostic code (department-level) |
| Flu-related hospitalizations | Weekly number of hospitalizations with a flu diagnosis | Hospitalization record (hospital-level) | Diagnostic code (department-level) |

\*ICU and ED stand for intensive care units and emergency departments, respectively.

### 3.4 Statistical analysis

Continuous variables were reported as medians with interquartile ranges (IQRs), and qualitative variables were reported as numbers and proportions (%). Quality and epidemiological indicators were calculated for the period from *t*_*init*_, a variable start date, to May 2022, a fixed end date. We chose this end date because of some technical issues affecting the integration of clinical reports into the CDW after this date. The temporal variations of quality indicators were modeled by linear functions:

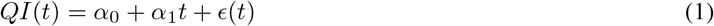

with *QI* the quality indicator, *t* the month, *α*_0_ and *α*_1_ parameters characterizing respectively the origin and the linear trend, and *ϵ*(*t*) a random error. We estimated model coefficients and 95% confidence intervals (CIs) using ordinary least squares regression. We discussed the linear trend *α*_1_ because it characterizes the temporal variation and could be affected by time-dependent data missingness. The temporal variation of the epidemiological indicators was discussed qualitatively, with particular emphasis on the post-COVID-19 period. Indeed, seasonal epidemics of bronchiolitis and flu were affected by COVID-19 and epidemiological indicators were used to adjust the response of healthcare organizations.

We performed these analyses using either a naive approach (N), which did not take into account the progressive digitalization of healthcare, or a novel approach, which we termed complete-source-only (CSO). To calculate the outcomes, we either used all available data (N) or restricted the analysis to healthcare sites (hospitals, departments, or units) where the EHR functionalities used to collect the required data were considered fully adopted before the start of the study period (CSO method, *t*_0_ ≤ *t*_*init*_ for each healthcare site and each data category used to calculate the outcome, as mentioned in Table 1). For the sake of simplicity, quality indicators were computed using the same denominator for both methods, considering cohorts of hospitalizations that occurred in the 28 of the 38 hospitals where hospitalization records were collected since January 2013 (i.e., with *t*_0_ for hospitalization records prior to that date).

We expected to observe indicators with increasing values when using the naive method, as the progressive adoption of EHR functionalities induces a temporally improving detection of outcomes. On the contrary, the CSO method aimed at stabilizing the data source used to detect outcomes to avoid this spurious temporal drift.

We conducted two sensitivity analyses to examine the quality indicators. First, we varied the value of *t*_*init*_ in [January 2013; January 2016; January 2019]. Second, we performed a subgroup analysis per hospital. Statistical analysis was performed using the Python package statmodels *v*0.13.5.[37] The modeling of EHR adoption was realized using the Python library EDS-TeVa *v*0.2.4 that has been made freely available.[38]

## 4 Results

### 4.1 Modeling of the EHR adoption

The APHP CDW contains data related to 14.5 million patients. The total amount of data available in the CDW increased over time, following dynamics that varied by data category and reflected the progressive digitization of healthcare (see Figure 1). While administrative records related to hospitalizations showed a stable collection for a decade, the other data categories showed a monotonic increase in collection, reflecting the progressive adoption of new EHR functionalities.

**Figure 1:**
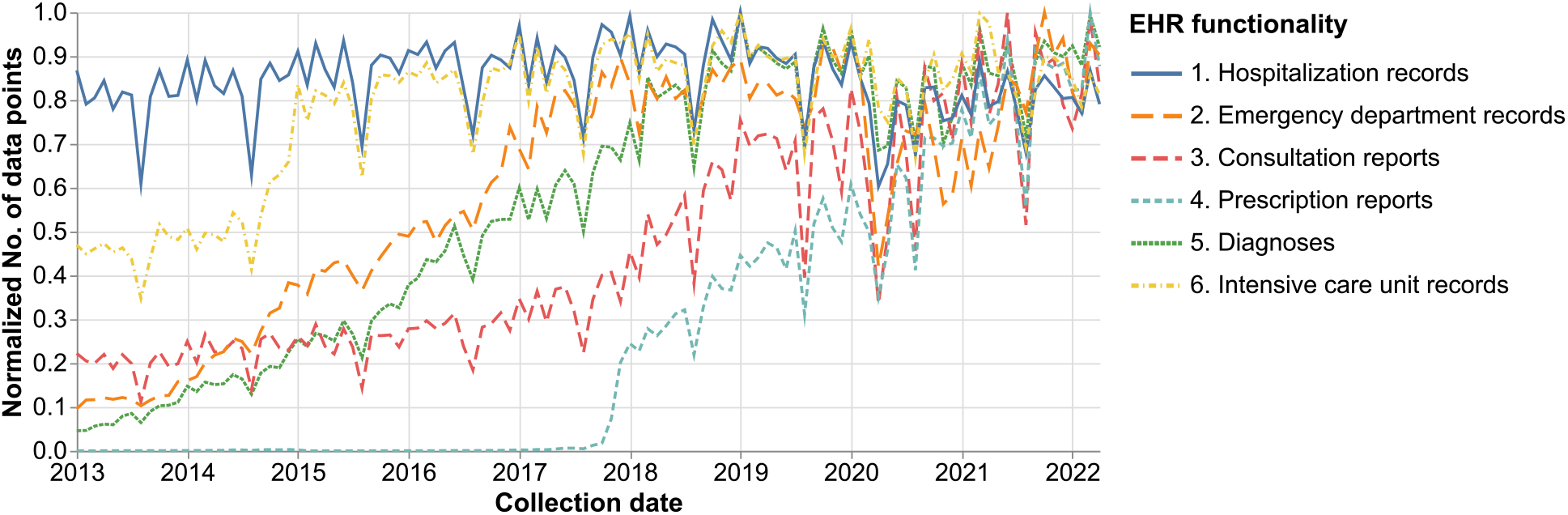
Temporal evolution of the amount of data collected within the electronic health record (EHR), per functionality. To obtain a common scale, for each functionality the monthly number of data points was divided by the highest value measured during the study period.

As shown in Figure2, we modeled the adoption of these functionalities in each healthcare site by fitting step functions with a level of description appropriate to the adoption mechanism (i.e., hospital, department, or unit). The digitization of hospitalization, ED, and ICU records was abrupt, whereas the adoption of EHR functionalities to capture prescription reports or consultation reports was more gradual. While the data availability curves per hospital and per department were mostly step-shaped, when we looked at the smaller unit level, here looking at ICU records, we observed adoptions that were rectangular (see Figure S1).

**Figure 2:**
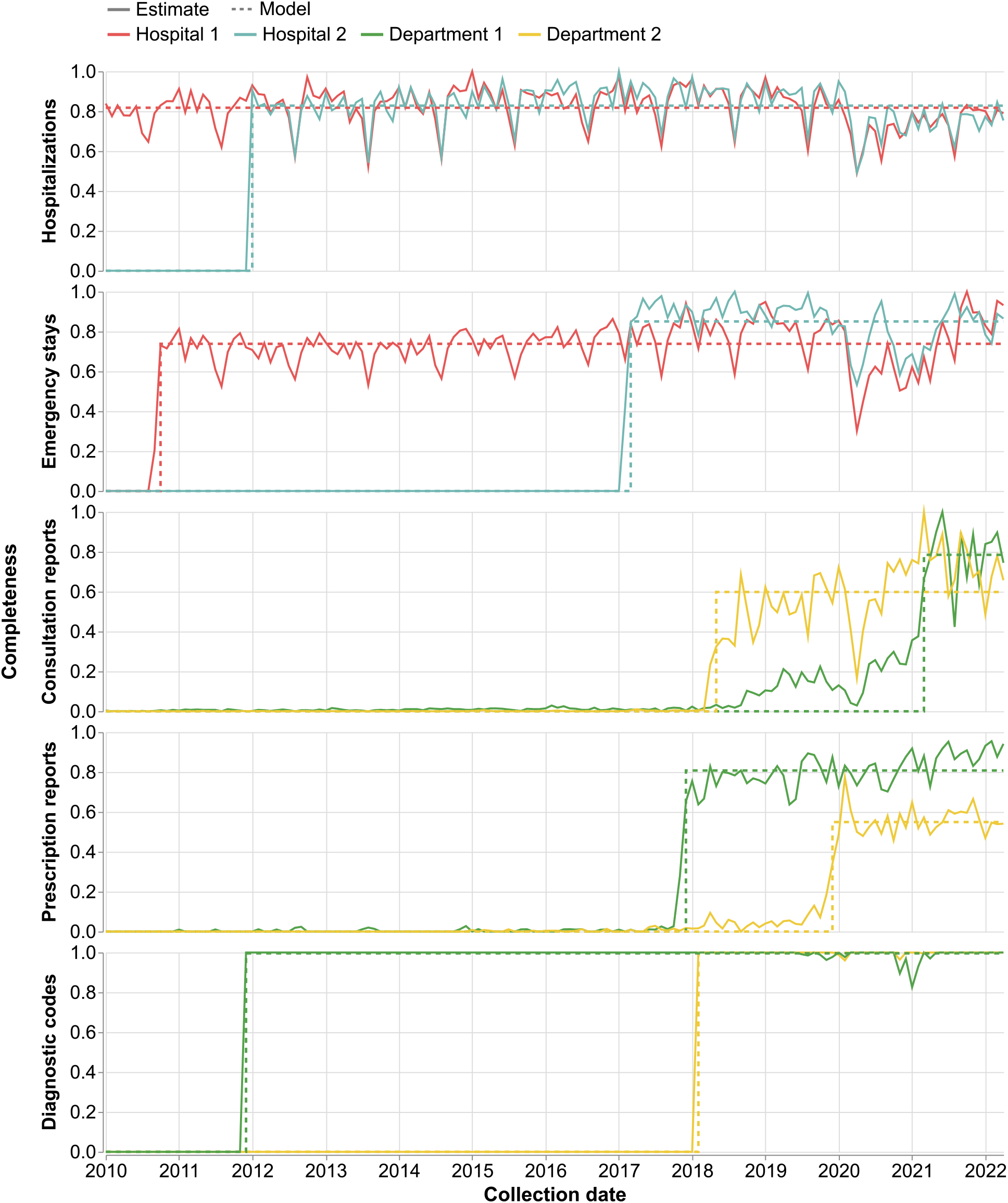
Completeness estimates (plain curves) of arbitrarily selected hospitals and departments considering from top to bottom hospitalization records, emergency department records, consultation reports, discharge prescriptions, and diagnostic codes. Department 1 and Department 2 are located within Hospital 1 and Hospital 2, respectively. Completeness estimate is defined either as the monthly number of data points divided by its maximum value during the study period (hospitalization and emergency department records, and consultation reports), or by the proportion of hospitalization records with at least one data point (diagnostic codes and prescription report). The modeling of the completeness estimate is shown as dashed curves.

### 4.2 Quality indicators

Figure 3 shows the temporal variations of the quality indicators using either the naive or the complete-source-only method, and choosing three different starting dates *t*_*init*_. Both methods resulted in similar curves for 30-day rehospitalization, but discrepancies appeared for other indicators. For 30-day ED consultation, 30-day consultation, and discharge prescription, a monotonic increase was observed with the naive method, which disappeared when the CSO method was applied. However, the application of the CSO method reduced the absolute value of the indicators, since the data used to determine the outcomes were filtered to obtain temporal stability of the data sources. The amplitude of this reduction was smaller for more recent starting dates. ICU readmission also showed a monotonic increase when using the naive method, but the application of the Step function CSO method resulted in a monotonically decreasing indicator (Figure S3). The use of a rectangular function CSO, which also takes into account the disappearance of units, resulted in a more stable indicator, as expected.

**Figure 3:**
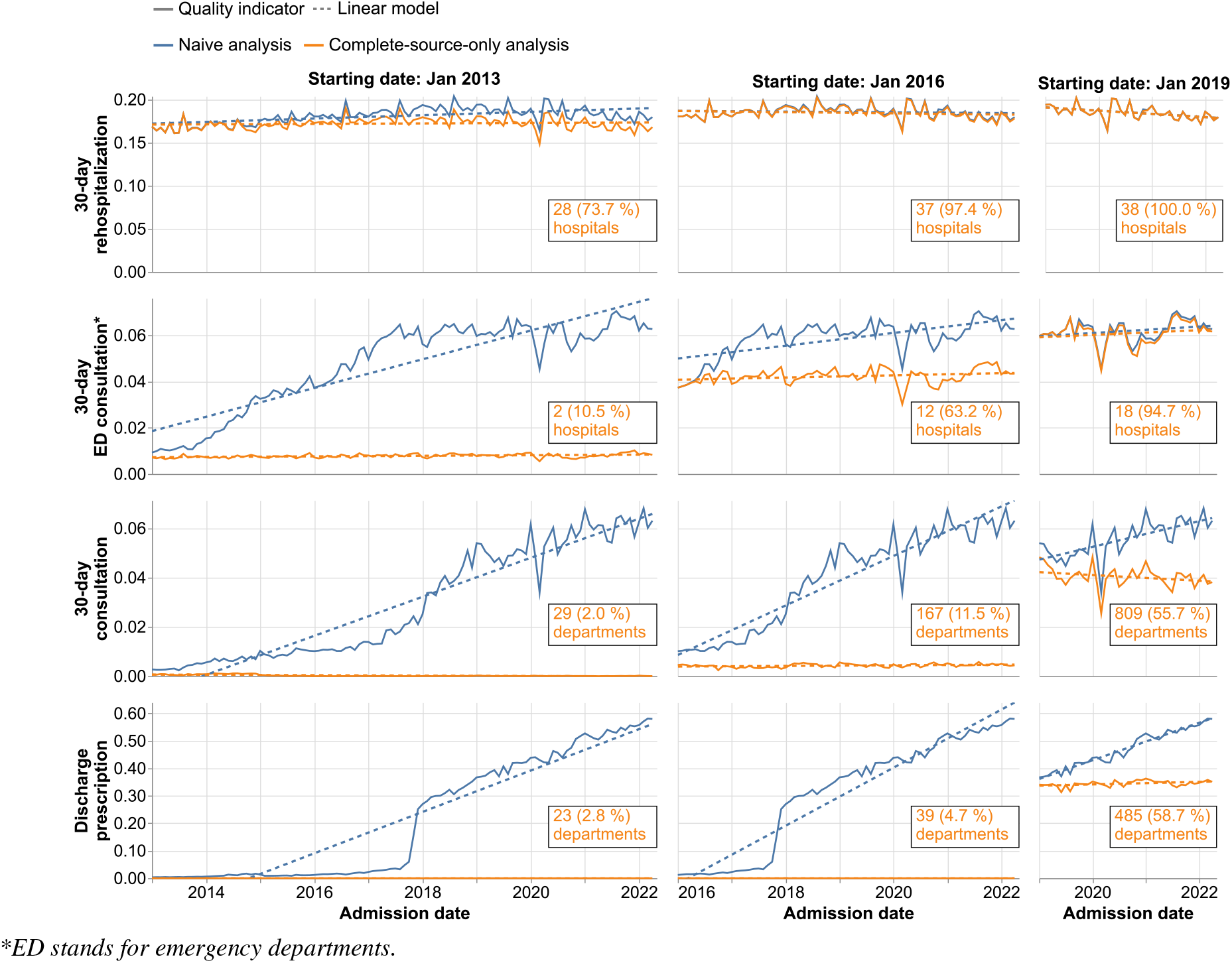
Combined effect of varying the initial observation date (*t*_*init*_) and the analysis (naive in blue and complete-source-only in orange) on the longitudinal study of quality indicators. The linear modeling of temporal variations is indicated by dashed lines. Insets indicate the number of healthcare sites selected in the complete-source-only analysis, along with the proportion they represent of healthcare sites with at least one data point collected by the EHR functionality during the study: January 2013 - May 2022.

As shown in the sensitivity analysis (Figure 4), the stabilizing effect of the CSO method was still observed when the 28 hospitals used for the cohort selection were considered separately. This sensitivity analysis also showed a strong reduction in the amplitude of the indicators induced by the CSO method, consistent with the observations in Figure 3. For 30-day ICU readmission, the sensitivity analysis on the 28 hospitals showed a positive slope for the naive analysis for older baseline data, a negative slope for the Step function CSO method, and no slope for the rectangular-function CSO method (Figure S4). Similar to the other quality indicators, this sensitivity analysis showed that both CSO methods reduced the amplitude of the indicator assessment. All fitted parameters are available in Tables S1 to S6 in the Supplementary Materials.

**Figure 4:**
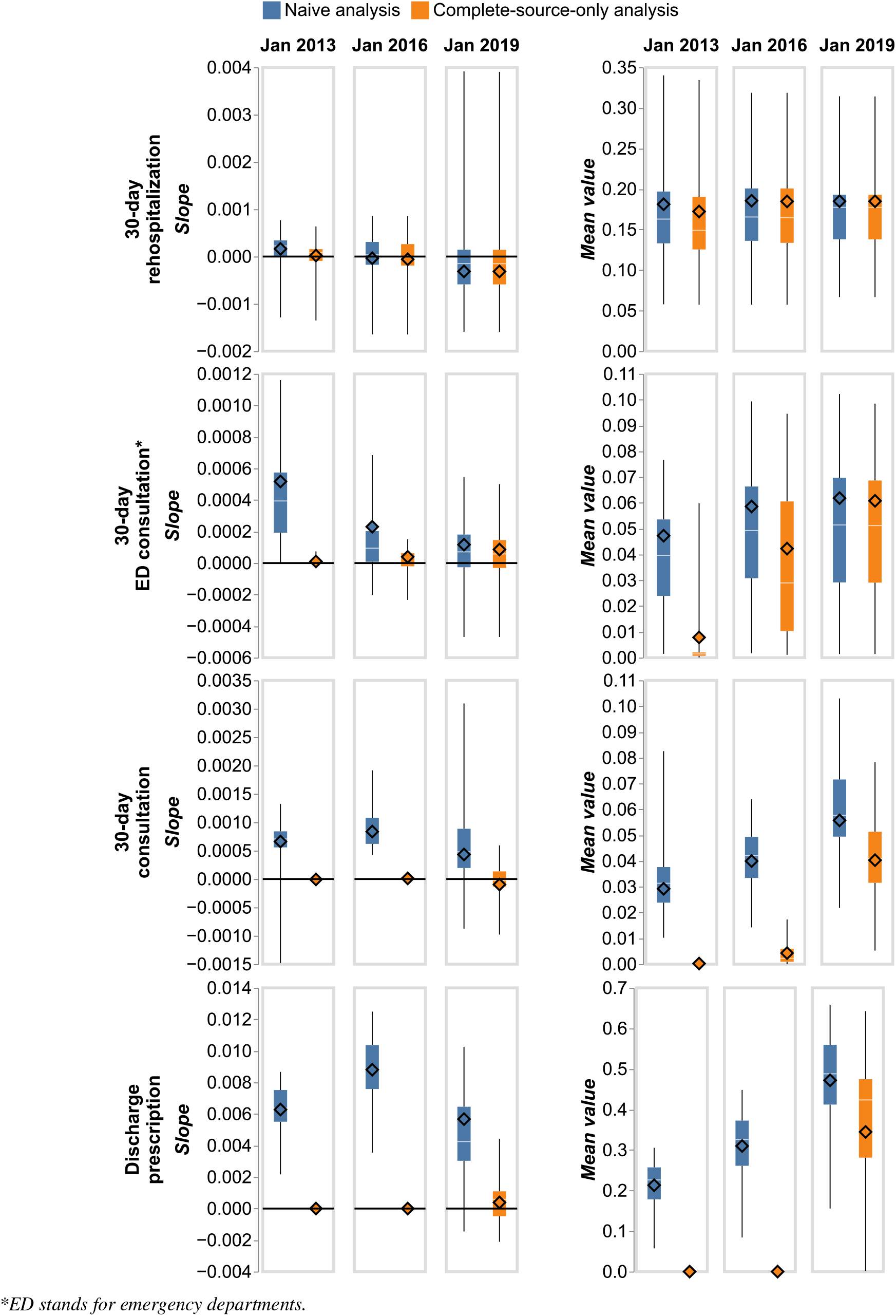
Left: slopes *α*_1_ of the linear model (Eq 1) estimated either on all the hospitals (diamond) or considering separately each one of the hospitals (IQR is the box, min and max are the lower and upper whiskers respectively), adopting either the naive (blue) or the complete-source-only (orange) method. Three different initial dates were considered. Right: values of the indicators averaged over the study period.

### 4.3 Epidemiological indicators

Figure 5 shows the weekly number of bronchiolitis- and flu-related hospitalizations estimated using either all diagnostic codes collected in the EHR (N), or only those collected in departments where the EHR functionality was fully adopted since the beginning of observation (CSO). While the strong impact of the COVID-19 context on these epidemics was observed in both cases, it seemed difficult to interpret the curves when using the naive method, as the COVID-19 effect could not be disentangled from the effect of the progressive EHR adoption. In fact, as can be seen by considering the years before the COVID-19 outbreak, this mechanism caused a spurious increase in the amplitude of the seasonal epidemics.

**Figure 5:**
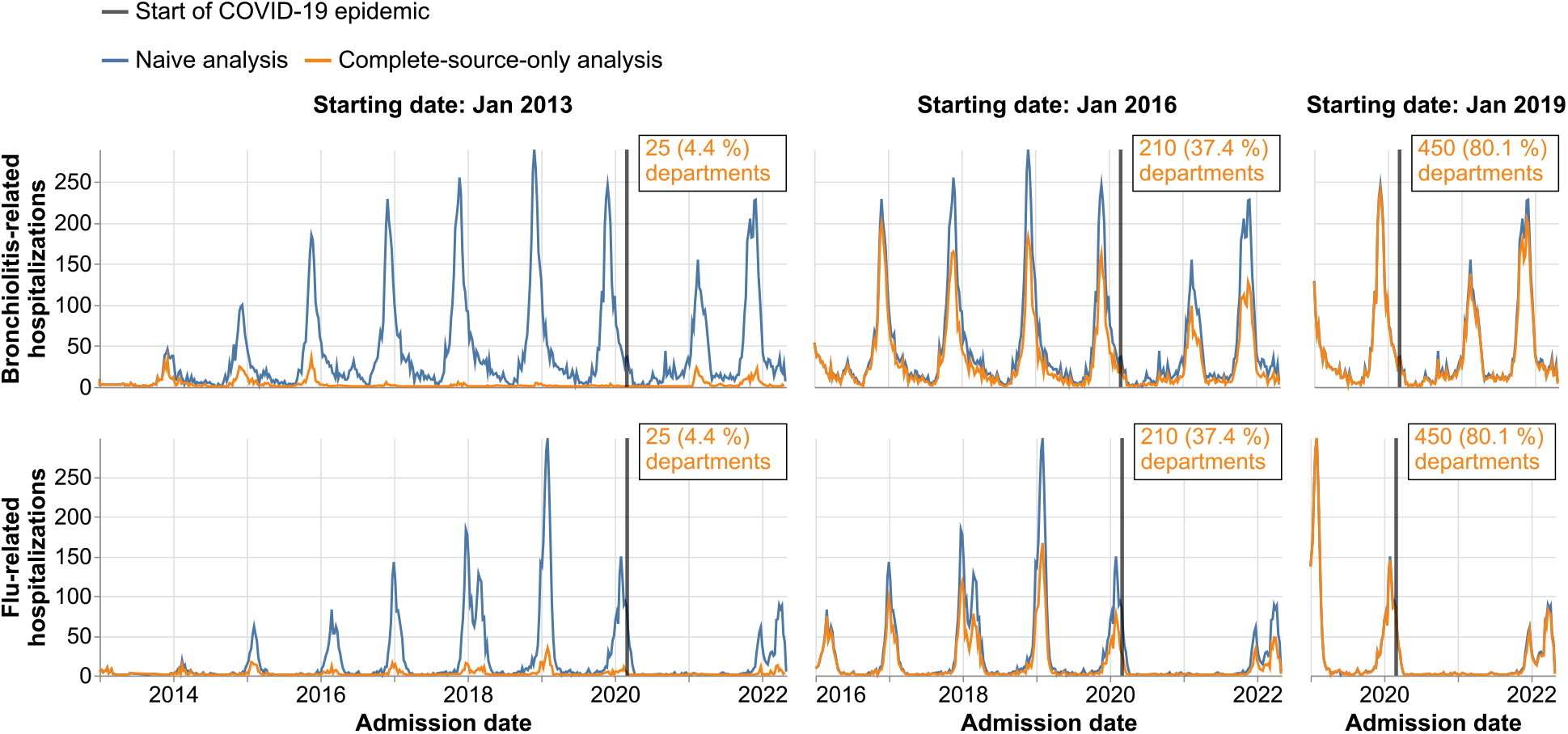
Epidemiological indicators computed adopting either the naive (blue) or the complete-source-only (orange) approach and varying the initial observation date (*t*_*init*_). The start of the COVID-19 epidemic is indicated by a gray line (March 2020). Insets indicate the number of healthcare sites selected in the complete-source-only analysis, along with the proportion they represent of healthcare sites with at least one data point collected by the EHR functionality during the study: January 2013 - May 2022.

## 5 Discussion

We showed that a multiscale modeling of digitization could be used to mitigate spurious temporal variations in the study of quality and epidemiological indicators. To this end, we discarded all healthcare sites that had not fully adopted the required EHR functionalities since the beginning of the measurement period, and referred to this approach as the complete-source-only method.

The time-dependent variation in completeness that we observed before any correction is consistent with previous work.[11] Our adjustment method is close to the well-known complete-case method, with the main difference being that we adopted a per-healthcare site approach instead of a per-case approach in order to leverage our understanding of the missingness mechanism induced by progressive digitization.[27] By taking advantage of a specific pattern that emerged when adopting this description, i.e., the abrupt increase or decrease in data availability, we were able to specifically adjust for the time-dependent bias induced by the progressive adoption of EHR functionalities, thus complementing other works that focused on the overall drifts or shifts of data distributions.[9, 10, 25] As discussed by Finlayson et al., many mechanisms can cause temporal variations, and we emphasize that this work addresses only one of them.[17]

The CSO method is not a panacea. On the one hand, its impact depends strongly on the time span examined. While too long a period leads to a massive discarding of data, too short a period can severely limit the scientific relevance of a longitudinal study. Therefore, an optimal trade-off should be found. Furthermore, this method is only useful when temporal drifts of statistical distributions are detrimental to the objective of the study. For example, it is not appropriate if the goal is instead to maximize the number of events/outcomes/patients detected while withstanding time-dependent biases. On the other hand, omitting data sources is not always harmless, as it may cause a selection bias by altering the population under study, e.g., omitting hospital units with patients with different severity. Consequently, the interpretation of results obtained using this strategy should remain cautious.

In addition to the specific advantages and limitations of the CSO method, this study illustrates some of the organizational challenges associated with real-world data platforms. Due to privacy concerns, investigators only have access to minimized cohorts of patients extracted from the full database which contains millions of records. However, the application of the analysis workflow presented in this study requires background information such as completeness estimated on the overall database. Such indicators should therefore be precomputed and provided to investigators in addition to patient-level data, requiring close coordination between platform operators and research teams. To address this challenge, we have structured the computer code of this project as an open source library that can be extended by investigators while being applied to the entire database by platform operators (see eFigure5 for the proposed workflow).

Our study has several limitations. First, we used a single, highly simplified definition of completeness. In some cases, it could be refined to match a more intuitive definition or a more elaborate definition of plausibility.[28] Second, we used a crude modeling of EHR adoption that relied on the detection of abrupt changes in data availability. The actual dynamics of a hospital’s information system exhibit diverse behaviors that may induce more complex patterns in statistical distributions.[39] Third, we considered highly simplified quality and epidemiological indicators. As such, the indicators we calculated are not directly applicable to epidemiological surveillance or quality monitoring. In particular, it seems important to further characterize the patients included in the analysis in order to be able to compare facilities with similar case mix.[32] Fourth, our analysis focused on some administrative and clinical data categories that do not cover the wide variety of data found in a CDW. It should therefore be expanded to support a larger range of studies conducted on real-world data.

## 6 Conclusion

EHR studies require working with data collected by a constantly evolving system. The current pace of technological innovation is unlikely to slow anytime soon, and this complexity will continue to increase. By focusing on a specific mechanism, the adoption of new EHR functionalities, we have shown that various metadata can be used to meaningfully analyze EHR data at scale. Moreover, automating the computation of such metadata was critical to avoid an explosion in data management burden. Our work is a step in the direction of developing tools and methods to address these challenges. Much work remains to be done, particularly to fully integrate information about technological change into the statistical design of studies.

## Author contributions statement

A.R., S.V and R.B. had full access to all the data in the study. They take responsibility for the integrity of the data and the accuracy of the data analysis.

- All authors designed the study.
- A.R and R.B. drafted the manuscript.
- All authors interpreted data and made critical intellectual revisions of the manuscript.
- R.B. did the literature review.
- A.R., B.P., P.B., S.V. developed the algorithms.
- R.B. supervised the project.

## Acknowledgments

We thank C. Taille, C. Chau and C. Baudoin for fruitful conversations, T. Petit-Jean for code proofreading and the clinical data warehouse of the Greater Paris University Hospitals for its support and the realization of data management and data curation tasks.

## Data availibility

Access to the clinical data warehouse’s raw data can be granted following the process described on its website: www.eds.aphp.fr. A prior validation of the access by the local institutional review board is required. In the case of non-AP-HP researchers, the signature of a collaboration contract is mandatory.

## Code availibility

The analyses of this article make extensive use of EDS-TeVa, a library developed in the context of AP-HP’s clinical data warehouse to automate the computation of indicators. It has been made publicly available under an open source license (BSD 3-clause): https://github.com/aphp/edsteva. A technical documentation of the library has been published to facilitate its use: https://aphp.github.io/edsteva/latest/. Moreover, the code developed to run the experiments of this study has also been made available freely in a separate github repository.[40]

## Supplementary Materials

### A Details on the modeling of EHR adoption

We used two alternative models of the adoption of elcetronic health record (EHR) functionalities: either a Step function model, which takes into account only the adoption of a functionality (most analyses, see Figure 2 in the main article), or a rectangular function model, which also takes into account the end of use of a functionality in a healthcare unit (complementary analyses related to the collection of intensive care unit -ICU-records, see Figure S1). This second modeling seems to be particularly appropriate when working at the unit level, since units are constantly appearing and disappearing due to hospital reorganizations.

**Figure S1:**
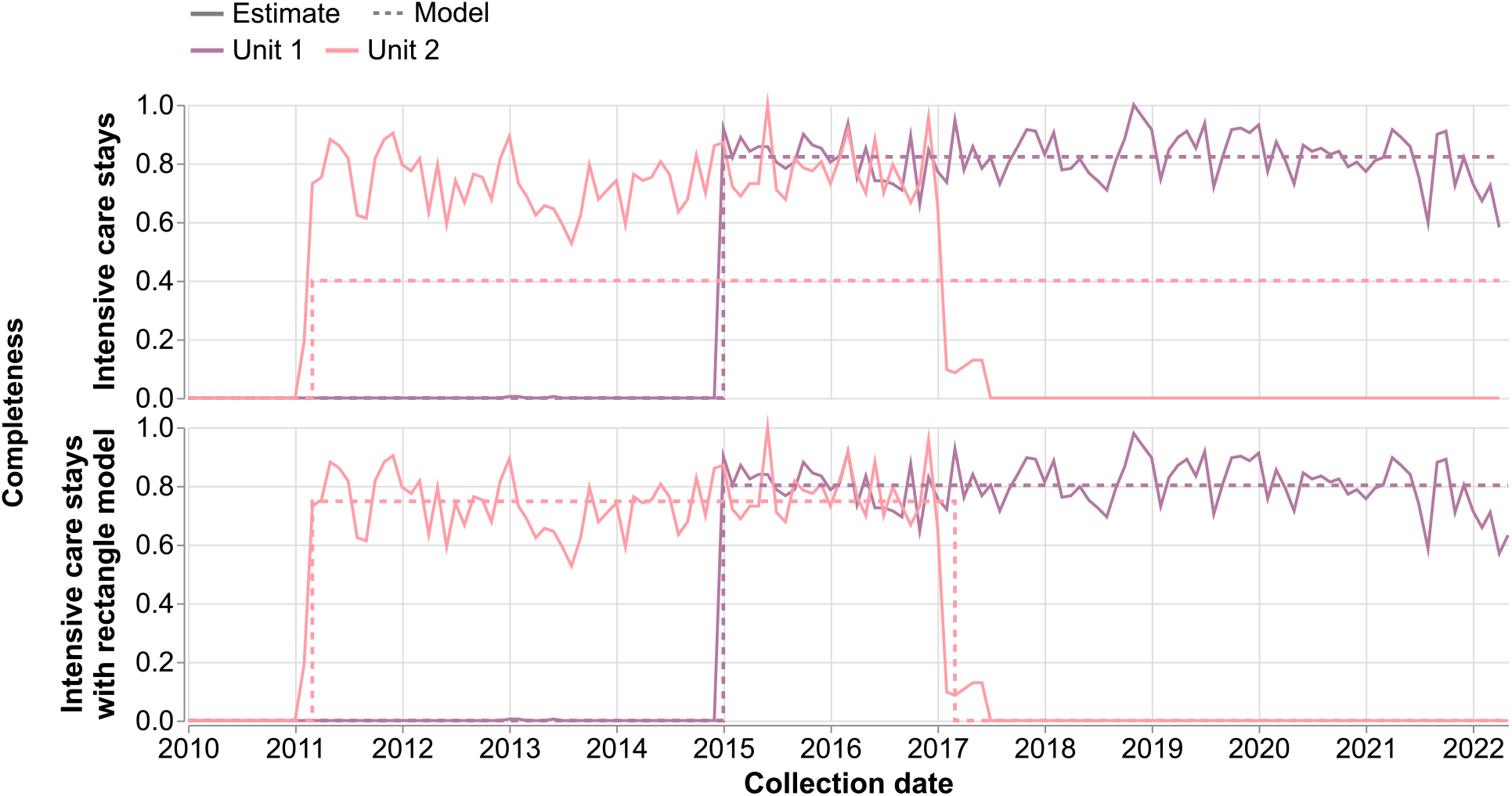
Completeness estimates of intensive care unit records (plain curve) considering two arbitrarily selected units (colors) along with their modeling (dashed lines) using either step functions (top) or rectangular functions (down). Completeness estimate is defined as the monthly number of data points divided by its maximum value during the study period.

#### A.1 Step function model

We used the following error definition to measure the distance between completeness estimates, *c*(*t*), and their modelings by step functions (*c*_0_ and *t*_0_ parameters):

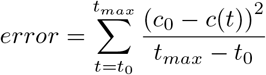

with *t*_*max*_ the end of the study period, i.e., May 2022. We underline that this definition leads to high values of error for healthcare sites that disappeared before the end of the study’s period.

#### A.2 Rectangular function model

We modified the fitted parameters and the definition of error when using rectangular functions to model EHR usage. In that case, in addition to *c*_0_ and *t*_0_ parameters the fit moreover provides *t*_1_, the end of functionality usage, and the error function is replaced by:

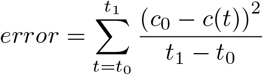

### B Goodness-of-fit of the Step function modeling

In the analyses presented in this article, we have not yet assessed the goodness-of-fit of our modeling. Figure S2 shows the dispersion of completeness estimates around the fitted Step function curve for each of the six data categories. Although the dispersion is not negligible, the pattern of an abrupt increase in data availability appears robust. Nevertheless, these curves show that the average value of the completeness estimate decreases after EHR adoption in the case of ICU records, consistent with the need to use rectangular functions as discussed earlier. For the sake of simplicity, we chose to remove healthcare sites with a low mean value of completeness after the estimated adoption date (*c*_0_ *≤* 0.15), which represent about 2% of the total number of administrative records, because their normalized completeness estimates would have spiked and made the figure unreadable.

**Figure S2:**
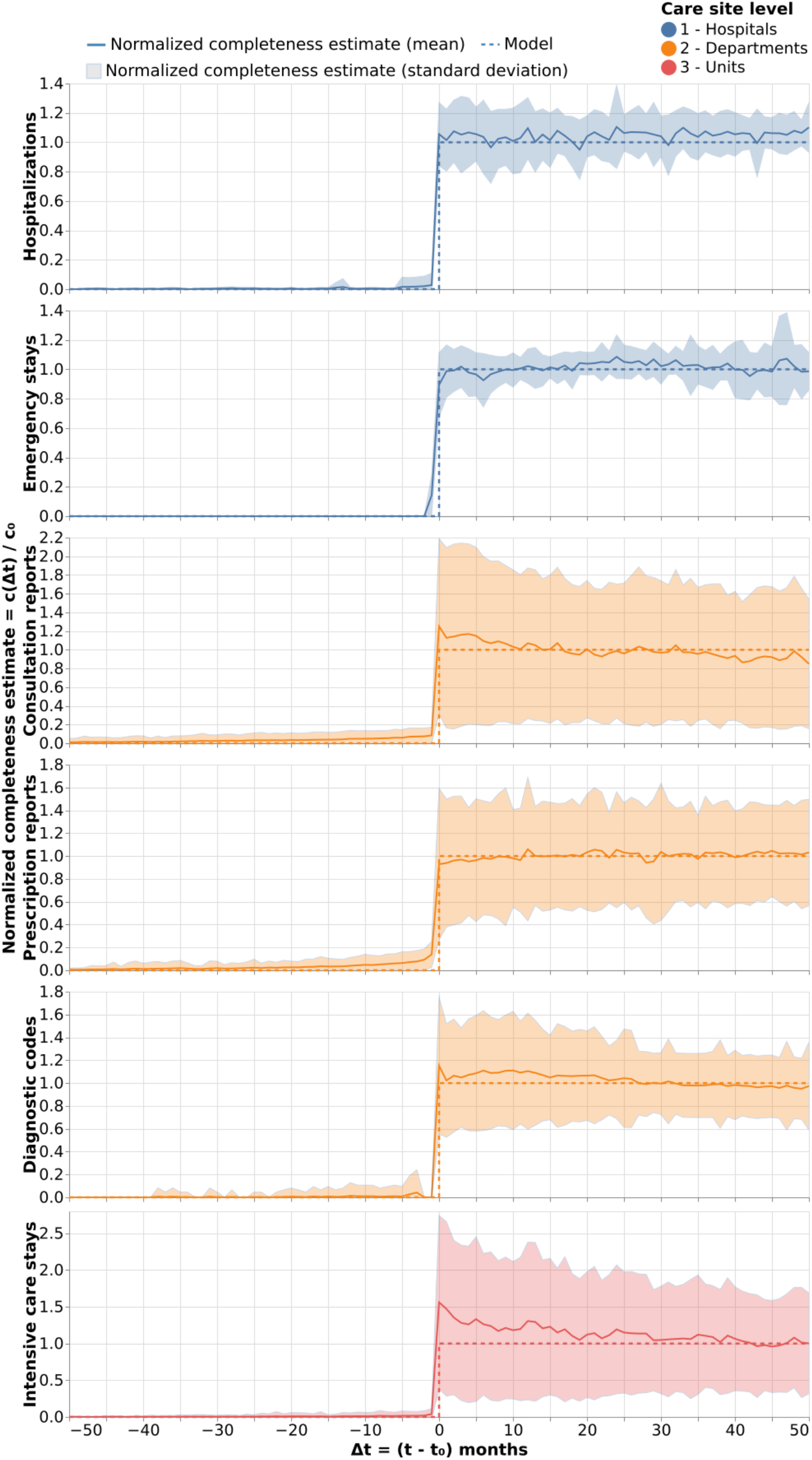
Completeness estimates centered on their adoption date *t*_0_ and normalized by their stabilized value *c*_0_, from top to bottom for each one of the six EHR functionalities and shown as their average (plain curve) and standard deviation (shaded area). The reference Step function is shown as dashed curves.

### C The special case of ICU-readmission

Since the model of ICU records is different from the other EHR functionalities, the analysis of 30-day ICU readmissions was treated separately. Figure S3 shows the temporal variation of the quality indicator using either the naive or the CSO method and choosing three different starting dates *t*_*init*_. The naive method resulted in a monotonous increase, while the Step function CSO method resulted in a monotonous decrease of the indicator. However, the rectangular function CSO, which also accounts for unit disappearance, discarded many small units and resulted in a stabilized indicator.

**Figure S3:**
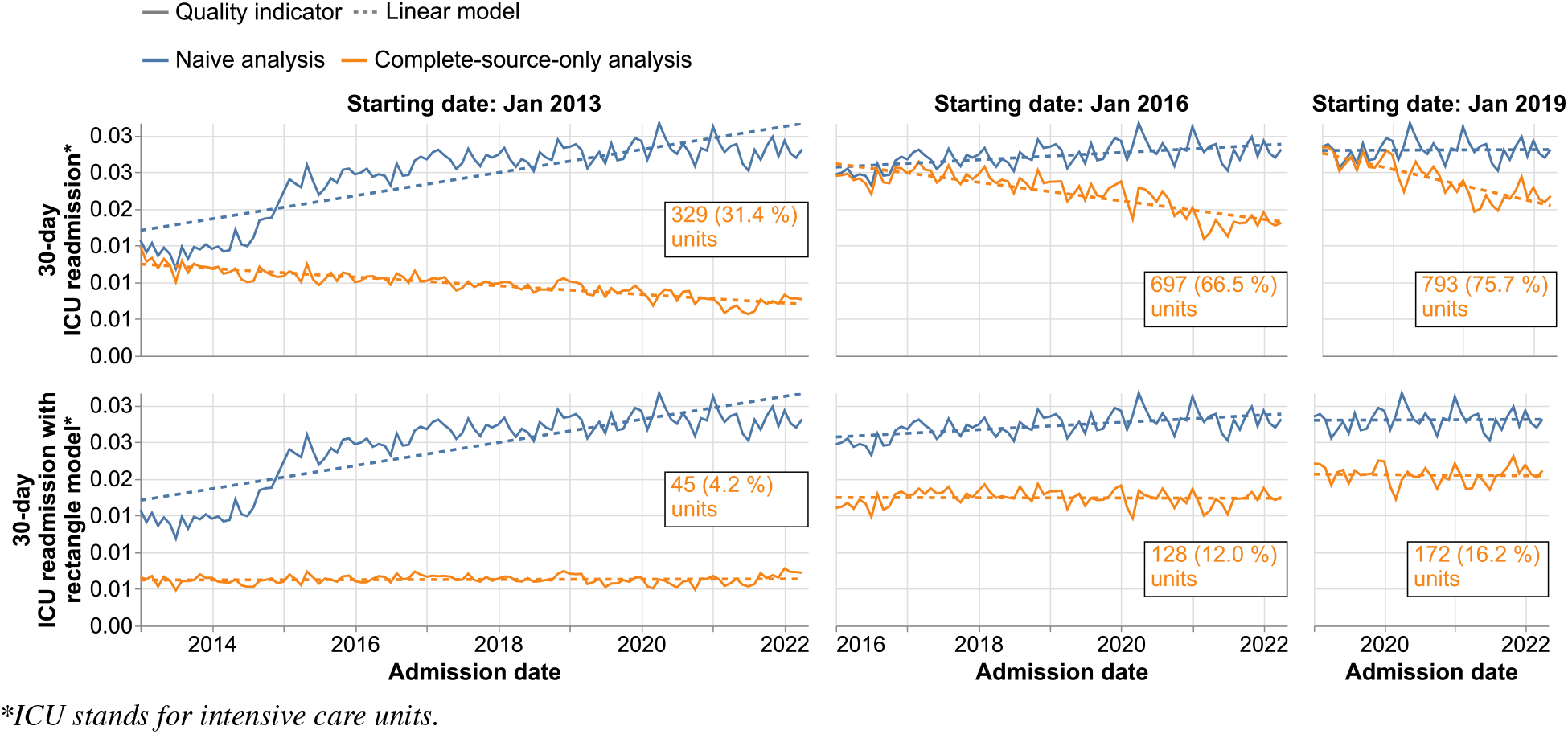
Combined effect of varying the initial observation date (*t*_*init*_) and the analysis (naive in blue and complete-source-only in orange) on the study of 30-day readmission in intensive care units. Two modelings of the adoption of the EHR functionality used to collect intensive care units records were compared, using either a step function (up) or a rectangular function (down). The linear modeling of temporal variations is indicated by dashed lines. Insets indicate the number of healthcare sites selected in the complete-source-only analysis, along with the proportion they represent of healthcare sites with at least one data point collected by the EHR functionality during the study: January 2013 - May 2022.

Figure S4 provides a sensitivity analysis of the slope *α*_1_ and the mean of the indicator for the 28 hospitals used for the cohort selection. Similar to the observations of Figure S3, it shows a positive slope for the naive analysis for older initial data, a negative slope for the step function CSO method, and no slope for the rectangular function CSO method. This sensitivity analysis also indicates that both CSO methods reduce the amplitude of the indicator.

**Figure S4:**
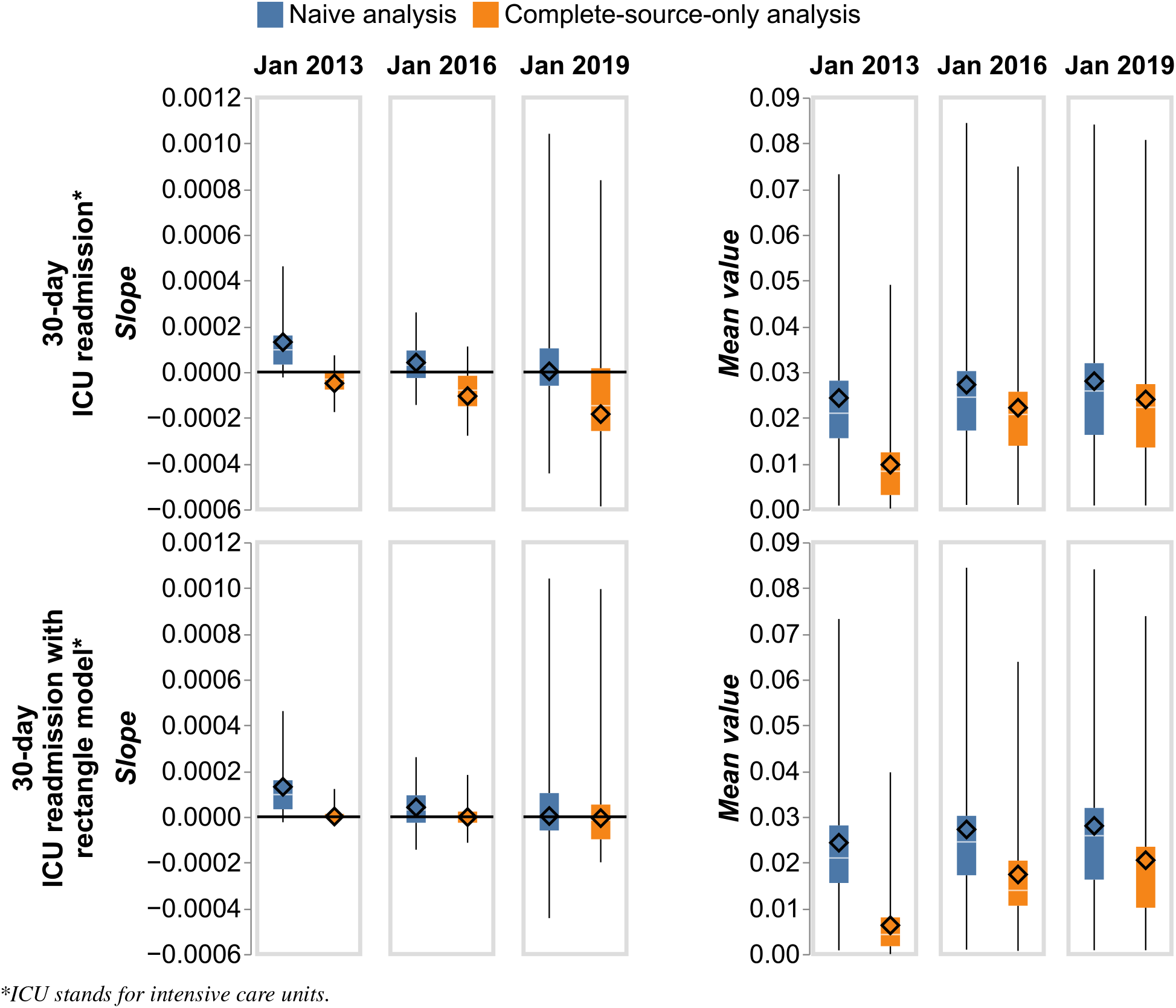
Left: slopes *α*_1_ of the linear model (Eq 1) estimated either on all the hospitals (diamond) or considering separately each one of the hospitals (IQR is the box, min and max are the lower and upper whiskers respectively), adopting either the naive (blue) or the complete-source-only (orange) method. Right: values of the indicators averaged over the study period. Two modelings of the adoption of intensive care units records were compared, using either a step function (up) or a rectangular function (down). Three different initial dates were considered.

### D Additional results

Tables S1 to S6 shows the detailed results of the modeling of quality and epidemiological indicators.

**Table S1:** Parameters resulting from the modeling of the temporal variations of the 30-day rehospitalization (origin *α*_0_ and slope *α*_1_, see Eq 1 in the main article) along with the number and proportion of considered hospitals for each method (naive and complete-source-only) and each start date.

| 30-day rehospitalization |  |  |  |  |
| --- | --- | --- | --- | --- |
| Start date | Statistical analysis | No. of hospitals considered in the analysis (%) | $\alpha_0$ [95%CI]* | $\alpha_1$ [95%CI]* |
| 2013-01-01 | Naive analysis | 38 (100 %) | $1.72 \cdot 10^{-1}$<br>[1.69·10<sup>-1</sup>, 1.75·10<sup>-1</sup>] | $1.62 \cdot 10^{-4}$<br>[1.16·10<sup>-4</sup>, 2.09·10<sup>-4</sup>] |
| | CSO analysis | 28 (73.7 %) | $1.71 \cdot 10^{-1}$<br>[1.68·10<sup>-1</sup>, 1.74·10<sup>-1</sup>] | $2.21 \cdot 10^{-5}$<br>[-1.72·10<sup>-5</sup>, 6.13·10<sup>-5</sup>] |
| 2016-01-01 | Naive analysis | 38 (100 %) | $1.87 \cdot 10^{-1}$<br>[1.84·10<sup>-1</sup>, 1.90·10<sup>-1</sup>] | $-3.55 \cdot 10^{-5}$<br>[-1.13·10<sup>-4</sup>, 4.19·10<sup>-5</sup>] |
| | CSO analysis | 37 (97.4 %) | $1.87 \cdot 10^{-1}$<br>[1.84·10<sup>-1</sup>, 1.90·10<sup>-1</sup>] | $-5.77 \cdot 10^{-5}$<br>[-1.34·10<sup>-4</sup>, 1.87·10<sup>-5</sup>] |
| 2019-01-01 | Naive analysis | 38 (100 %) | $1.91 \cdot 10^{-1}$<br>[1.86·10<sup>-1</sup>, 1.96·10<sup>-1</sup>] | $-3.13 \cdot 10^{-4}$<br>[-5.17·10<sup>-4</sup>, -1.10·10<sup>-4</sup>] |
| | CSO analysis | 38 (100.0 %) | $1.91 \cdot 10^{-1}$<br>[1.86·10<sup>-1</sup>, 1.96·10<sup>-1</sup>] | $-3.15 \cdot 10^{-4}$<br>[-5.17·10<sup>-4</sup>, -1.13·10<sup>-4</sup>] |

\*The confidence interval is based on Student's *t*-distribution.

**Table S2:**
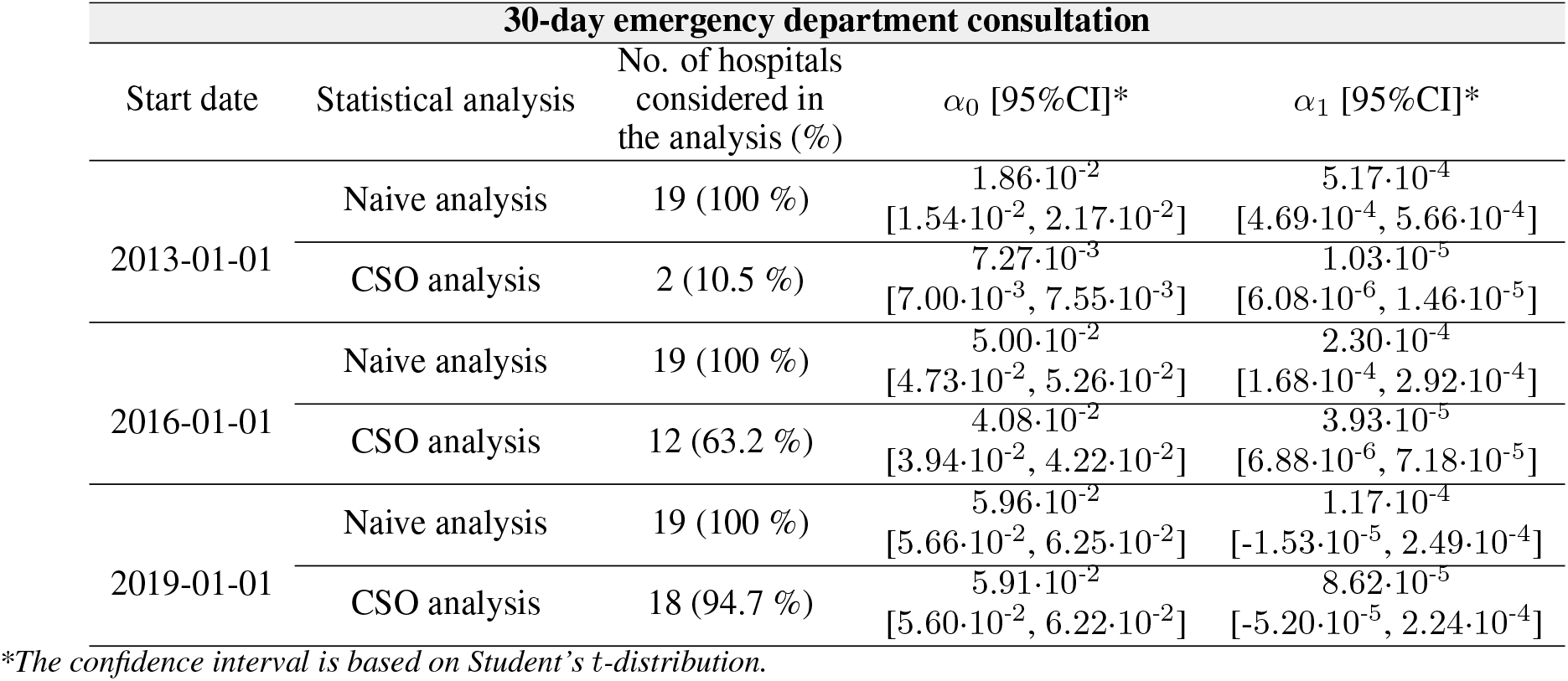
Parameters resulting from the modeling of the temporal variations of the 30-day emergency departments consultation (origin *α*_0_ and slope *α*_1_, see Eq 1 in the main article) along with the number and proportion of considered hospitals for each method (naive and complete-source-only) and each start date.

| 30-day emergency department consultation |  |  |  |  |
| --- | --- | --- | --- | --- |
| Start date | Statistical analysis | No. of hospitals considered in the analysis (%) | $\alpha_0$ [95%CI]* | $\alpha_1$ [95%CI]* |
| 2013-01-01 | Naive analysis | 19 (100 %) | $1.86 \cdot 10^{-2}$<br>[1.54·10<sup>-2</sup>, 2.17·10<sup>-2</sup>] | $5.17 \cdot 10^{-4}$<br>[4.69·10<sup>-4</sup>, 5.66·10<sup>-4</sup>] |
| | CSO analysis | 2 (10.5 %) | $7.27 \cdot 10^{-3}$<br>[7.00·10<sup>-3</sup>, 7.55·10<sup>-3</sup>] | $1.03 \cdot 10^{-5}$<br>[6.08·10<sup>-6</sup>, 1.46·10<sup>-5</sup>] |
| 2016-01-01 | Naive analysis | 19 (100 %) | $5.00 \cdot 10^{-2}$<br>[4.73·10<sup>-2</sup>, 5.26·10<sup>-2</sup>] | $2.30 \cdot 10^{-4}$<br>[1.68·10<sup>-4</sup>, 2.92·10<sup>-4</sup>] |
| | CSO analysis | 12 (63.2 %) | $4.08 \cdot 10^{-2}$<br>[3.94·10<sup>-2</sup>, 4.22·10<sup>-2</sup>] | $3.93 \cdot 10^{-5}$<br>[6.88·10<sup>-6</sup>, 7.18·10<sup>-5</sup>] |
| 2019-01-01 | Naive analysis | 19 (100 %) | $5.96 \cdot 10^{-2}$<br>[5.66·10<sup>-2</sup>, 6.25·10<sup>-2</sup>] | $1.17 \cdot 10^{-4}$<br>[-1.53·10<sup>-5</sup>, 2.49·10<sup>-4</sup>] |
| | CSO analysis | 18 (94.7 %) | $5.91 \cdot 10^{-2}$<br>[5.60·10<sup>-2</sup>, 6.22·10<sup>-2</sup>] | $8.62 \cdot 10^{-5}$<br>[-5.20·10<sup>-5</sup>, 2.24·10<sup>-4</sup>] |

\*The confidence interval is based on Student's *t*-distribution.

**Table S3:** Parameters resulting from the modeling of the temporal variations of the 30-day consultation (origin *α*_0_ and slope *α*_1_, see Eq 1 in the main article) along with the number and proportion of considered departments for each method (naive and complete-source-only) and each start date.

| 30-day consultation |  |  |  |  |
| --- | --- | --- | --- | --- |
| Start date | Statistical analysis | No. of departments considered in the analysis (%) | $\alpha_0$ [95%CI]* | $\alpha_1$ [95%CI]* |
| 2013-01-01 | Naive analysis | 1452 (100 %) | $-7.72 \cdot 10^{-3}$<br>[-1.03·10<sup>-2</sup>, -5.13·10<sup>-3</sup>] | $6.72 \cdot 10^{-4}$<br>[6.32·10<sup>-4</sup>, 7.12·10<sup>-4</sup>] |
| | CSO analysis | 29 (2.0 %) | $6.12 \cdot 10^{-4}$<br>[5.17·10<sup>-4</sup>, 7.06·10<sup>-4</sup>] | $-6.80 \cdot 10^{-6}$<br>[-8.27·10<sup>-6</sup>, -5.33·10<sup>-6</sup>] |
| 2016-01-01 | Naive analysis | 1452 (100 %) | $8.27 \cdot 10^{-3}$<br>[5.40·10<sup>-3</sup>, 1.11·10<sup>-2</sup>] | $8.58 \cdot 10^{-4}$<br>[7.92·10<sup>-4</sup>, 9.24·10<sup>-4</sup>] |
| | CSO analysis | 167 (11.5 %) | $3.86 \cdot 10^{-3}$<br>[3.55·10<sup>-3</sup>, 4.16·10<sup>-3</sup>] | $1.38 \cdot 10^{-5}$<br>[6.79·10<sup>-6</sup>, 2.07·10<sup>-5</sup>] |
| 2019-01-01 | Naive analysis | 1452 (100 %) | $4.70 \cdot 10^{-2}$<br>[4.35·10<sup>-2</sup>, 5.05·10<sup>-2</sup>] | $4.89 \cdot 10^{-4}$<br>[3.33·10<sup>-4</sup>, 6.45·10<sup>-4</sup>] |
| | CSO analysis | 809 (55.7 %) | $4.21 \cdot 10^{-2}$<br>[3.94·10<sup>-2</sup>, 4.47·10<sup>-2</sup>] | $-6.78 \cdot 10^{-5}$<br>[-1.84·10<sup>-4</sup>, 4.87·10<sup>-5</sup>] |

\*The confidence interval is based on Student's *t*-distribution.

**Table S4:** Parameters resulting from the modeling of the temporal variations of the discharge prescription (origin *α*_0_ and slope *α*_1_, see Eq 1 in the main article) along with the number and proportion of considered departments for each method (naive and complete-source-only) and each start date.

| Discharge prescription |  |  |  |  |
| --- | --- | --- | --- | --- |
| Start date | Statistical analysis | No. of departments considered in the analysis (%) | $\alpha_0$ [95%CI]* | $\alpha_1$ [95%CI]* |
| 2013-01-01 | Naive analysis | 825 (100 %) | $-1.36 \cdot 10^{-1}$<br>[-1.64·10<sup>-1</sup>, -1.07·10<sup>-1</sup>] | $6.28 \cdot 10^{-3}$<br>[5.83·10<sup>-3</sup>, 6.73·10<sup>-3</sup>] |
| | CSO analysis | 23 (2.8 %) | $7.97 \cdot 10^{-5}$<br>[6.53·10<sup>-5</sup>, 9.41·10<sup>-5</sup>] | $-1.00 \cdot 10^{-6}$<br>[-1.22·10<sup>-6</sup>, -7.75·10<sup>-7</sup>] |
| 2016-01-01 | Naive analysis | 825 (100 %) | $-2.03 \cdot 10^{-2}$<br>[-4.52·10<sup>-2</sup>, 4.65·10<sup>-3</sup>] | $8.81 \cdot 10^{-3}$<br>[8.23·10<sup>-3</sup>, 9.38·10<sup>-3</sup>] |
| | CSO analysis | 39 (4.7 %) | $3.19 \cdot 10^{-5}$<br>[1.57·10<sup>-5</sup>, 4.82·10<sup>-5</sup>] | $2.15 \cdot 10^{-7}$<br>[-1.59·10<sup>-7</sup>, 5.89·10<sup>-7</sup>] |
| 2019-01-01 | Naive analysis | 825 (100 %) | $3.62 \cdot 10^{-1}$<br>[3.53·10<sup>-1</sup>, 3.70·10<sup>-1</sup>] | $5.66 \cdot 10^{-3}$<br>[5.28·10<sup>-3</sup>, 6.04·10<sup>-3</sup>] |
| | CSO analysis | 485 (58.7 %) | $3.37 \cdot 10^{-1}$<br>[3.31·10<sup>-1</sup>, 3.44·10<sup>-1</sup>] | $3.67 \cdot 10^{-4}$<br>[9.47·10<sup>-5</sup>, 6.40·10<sup>-4</sup>] |

\*The confidence interval is based on Student's *t*-distribution.

**Table S5:** Parameters resulting from the modeling of the temporal variations of the 30-day intensive care units readmission (origin *α*_0_ and slope *α*_1_, see Eq 1 in the main article) along with the number and proportion of considered units for each method (naive, step function complete-source-only and rectangular function complete-source-only) and each start date.

| **30-day intensive care unit readmission** |  |  |  |  |
| --- | --- | --- | --- | --- |
| Start date | Statistical analysis | No. of units considered in the analysis (%) | $\alpha_0$ [95%CI]* | $\alpha_1$ [95%CI]* |
| 2013-01-01 | Naive analysis | 1048 (100 %) | $1.71 \cdot 10^{-2}$<br>[$1.60 \cdot 10^{-2}$, $1.81 \cdot 10^{-2}$] | $1.31 \cdot 10^{-4}$<br>[$1.15 \cdot 10^{-4}$, $1.47 \cdot 10^{-4}$] |
| | Step function CSO | 329 (31.4 %) | $1.25 \cdot 10^{-2}$<br>[$1.22 \cdot 10^{-2}$, $1.28 \cdot 10^{-2}$] | $-4.94 \cdot 10^{-5}$<br>[$-5.41 \cdot 10^{-5}$, $-4.48 \cdot 10^{-5}$] |
| | Rectangular function CSO | 45 (4.2 %) | $6.21 \cdot 10^{-3}$<br>[$6.00 \cdot 10^{-3}$, $6.43 \cdot 10^{-3}$] | $1.09 \cdot 10^{-6}$<br>[$-2.25 \cdot 10^{-6}$, $4.43 \cdot 10^{-6}$] |
| 2016-01-01 | Naive analysis | 1048 (100 %) | $2.57 \cdot 10^{-2}$<br>[$2.51 \cdot 10^{-2}$, $2.63 \cdot 10^{-2}$] | $4.16 \cdot 10^{-5}$<br>[$2.77 \cdot 10^{-5}$, $5.56 \cdot 10^{-5}$] |
| | Step function CSO | 697 (66.5 %) | $2.61 \cdot 10^{-2}$<br>[$2.55 \cdot 10^{-2}$, $2.68 \cdot 10^{-2}$] | $-1.05 \cdot 10^{-4}$<br>[$-1.20 \cdot 10^{-4}$, $-9.04 \cdot 10^{-5}$] |
| | Rectangular function CSO | 128 (12.0 %) | $1.75 \cdot 10^{-2}$<br>[$1.70 \cdot 10^{-2}$, $1.80 \cdot 10^{-2}$] | $-2.32 \cdot 10^{-6}$<br>[$-1.33 \cdot 10^{-5}$, $8.62 \cdot 10^{-6}$] |
| 2019-01-01 | Naive analysis | 1048 (100 %) | $2.80 \cdot 10^{-2}$<br>[$2.71 \cdot 10^{-2}$, $2.88 \cdot 10^{-2}$] | $3.32 \cdot 10^{-6}$<br>[$-3.64 \cdot 10^{-5}$, $4.30 \cdot 10^{-5}$] |
| | Step function CSO | 793 (75.7 %) | $2.76 \cdot 10^{-2}$<br>[$2.67 \cdot 10^{-2}$, $2.85 \cdot 10^{-2}$] | $-1.84 \cdot 10^{-4}$<br>[$-2.25 \cdot 10^{-4}$, $-1.44 \cdot 10^{-4}$] |
| | Rectangular function CSO | 172 (16.2 %) | $2.06 \cdot 10^{-2}$<br>[$1.98 \cdot 10^{-2}$, $2.14 \cdot 10^{-2}$] | $-4.64 \cdot 10^{-6}$<br>[$-3.94 \cdot 10^{-5}$, $3.02 \cdot 10^{-5}$] |

\*The confidence interval is based on Student's *t*-distribution.

**Table S6:** Number of departments considered for the computation of the epidemiological indicators for each method and each start date.

| **Bronchiolitis and Flu-related hospitalizations** |  |  |
| --- | --- | --- |
| Start date | Statistical analysis | No. of departments considered in the analysis (%) |
| 2013-01-01 | Naive analysis | 562 (100 %) |
|  | CSO analysis | 25 (4.4 %) |
| 2016-01-01 | Naive analysis | 562 (100 %) |
|  | CSO analysis | 210 (37.4 %) |
| 2019-01-01 | Naive analysis | 562 (100 %) |
|  | CSO analysis | 450 (80.1 %) |

### E Delivery of indicators in the context of a clinical data warehouse

The indicators used in this study’s analyses were computed on the entire database of the clinical data warehouse, to which access is never granted to investigators that are not members of the IT department in charge of the platform operation (i.e., data custodian) in order to protect patient privacy (data minimization principle). In fact, investigators analyze data extracts related to specific cohorts that are provided by data custodians in isolated and secure environments (see Figure S5). The provision of pre-computed indicators describing EHR adoption, in addition to patient-level data, therefore appears necessary for investigators to use the complete-source-only method.

**Figure S5:**
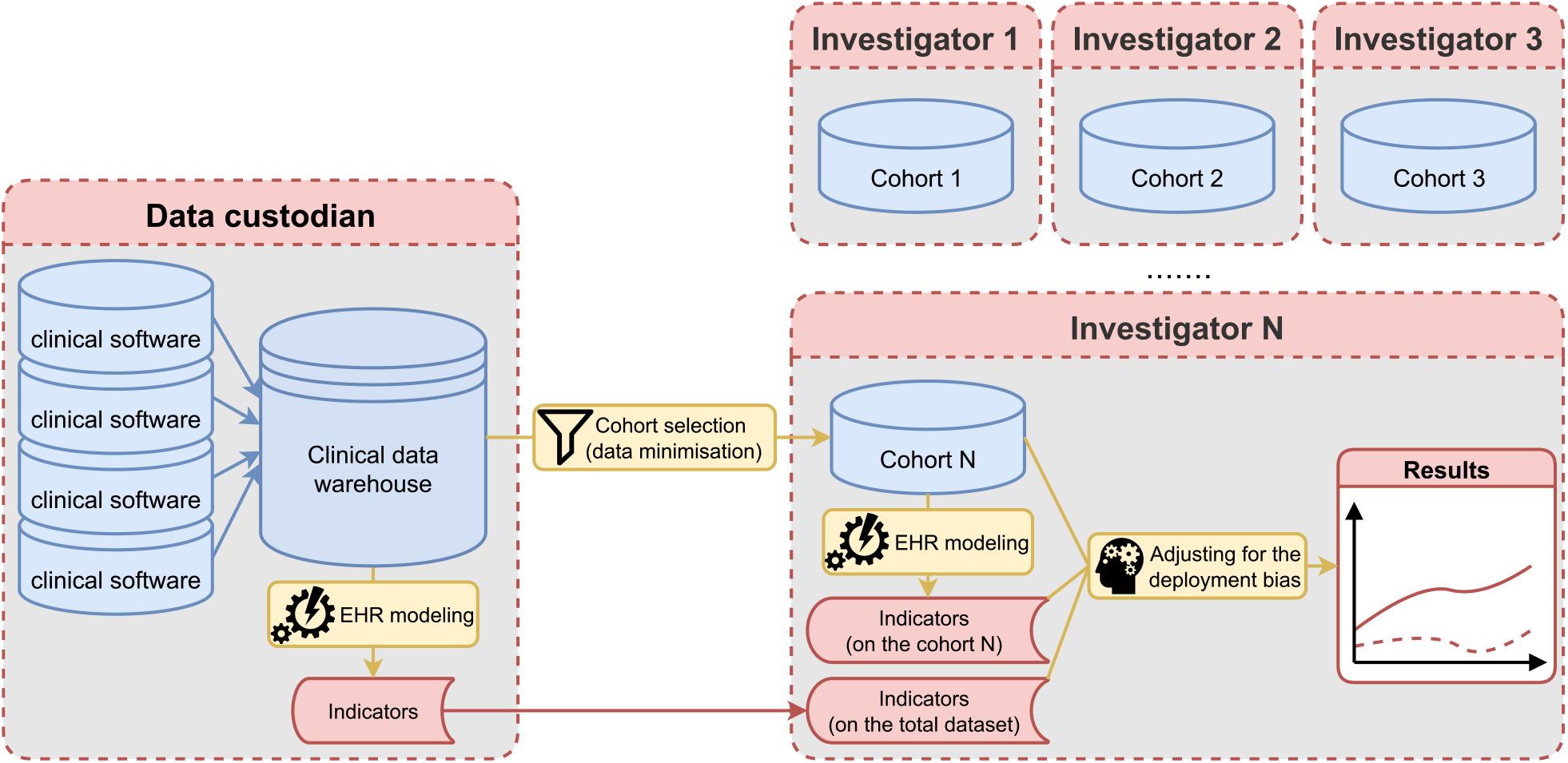
Data flows in the context of a clinical data warehouse. Blue: from left to right, patient-level data collected in clinical softwares are integrated and standardized by data custodians and data relative to cohorts of patients are extracted to be analyzed by investigators in per-project isolated environments. Red: in order to adjust for the progressive EHR adoption, dedicated indicators are computed either by investigators on their cohorts or by data custodians on the total dataset. Data custodians’ and investigators’ data accesses are represented by dashed red boxes.

### F The RECORD’s statement checklist

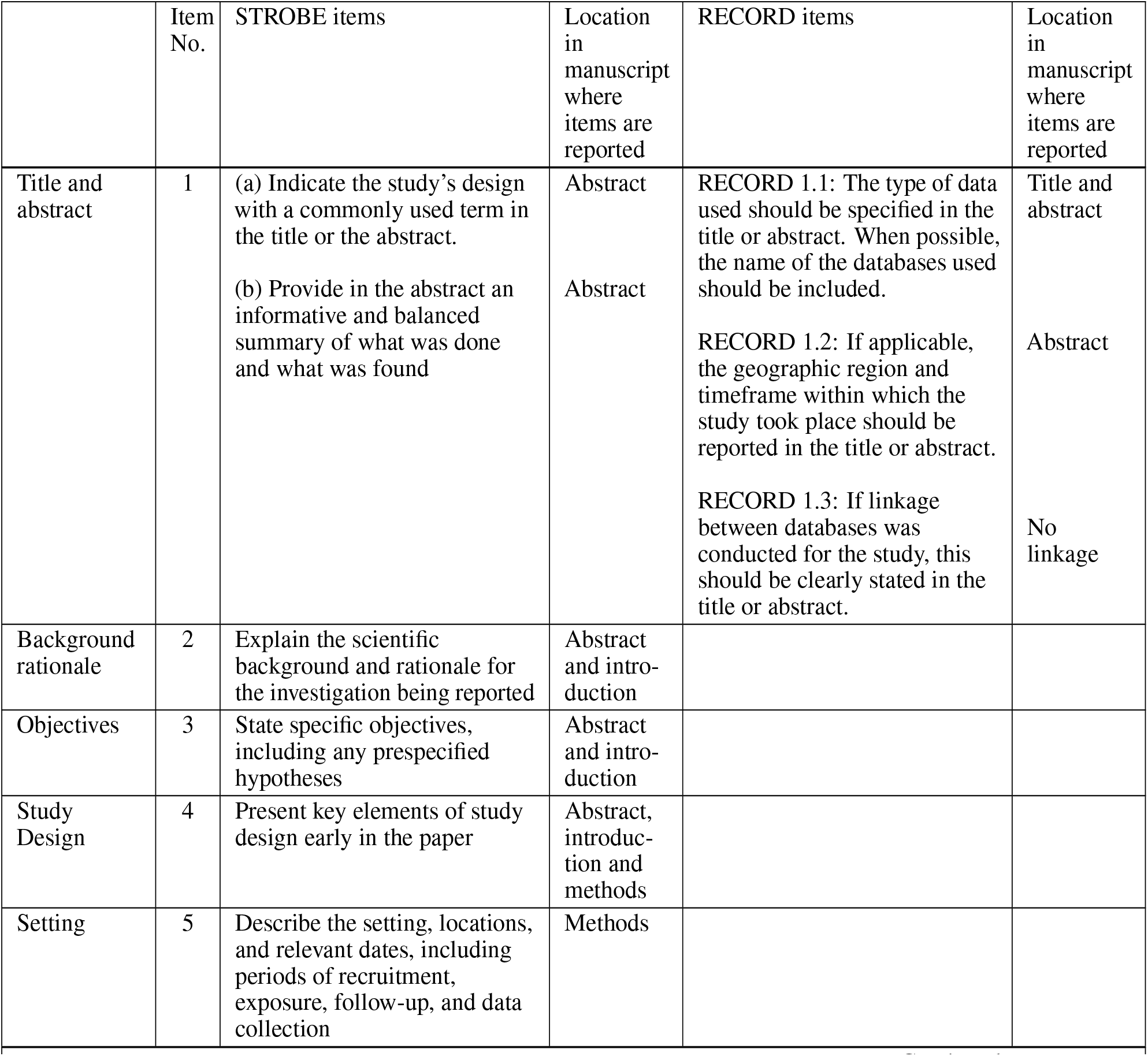
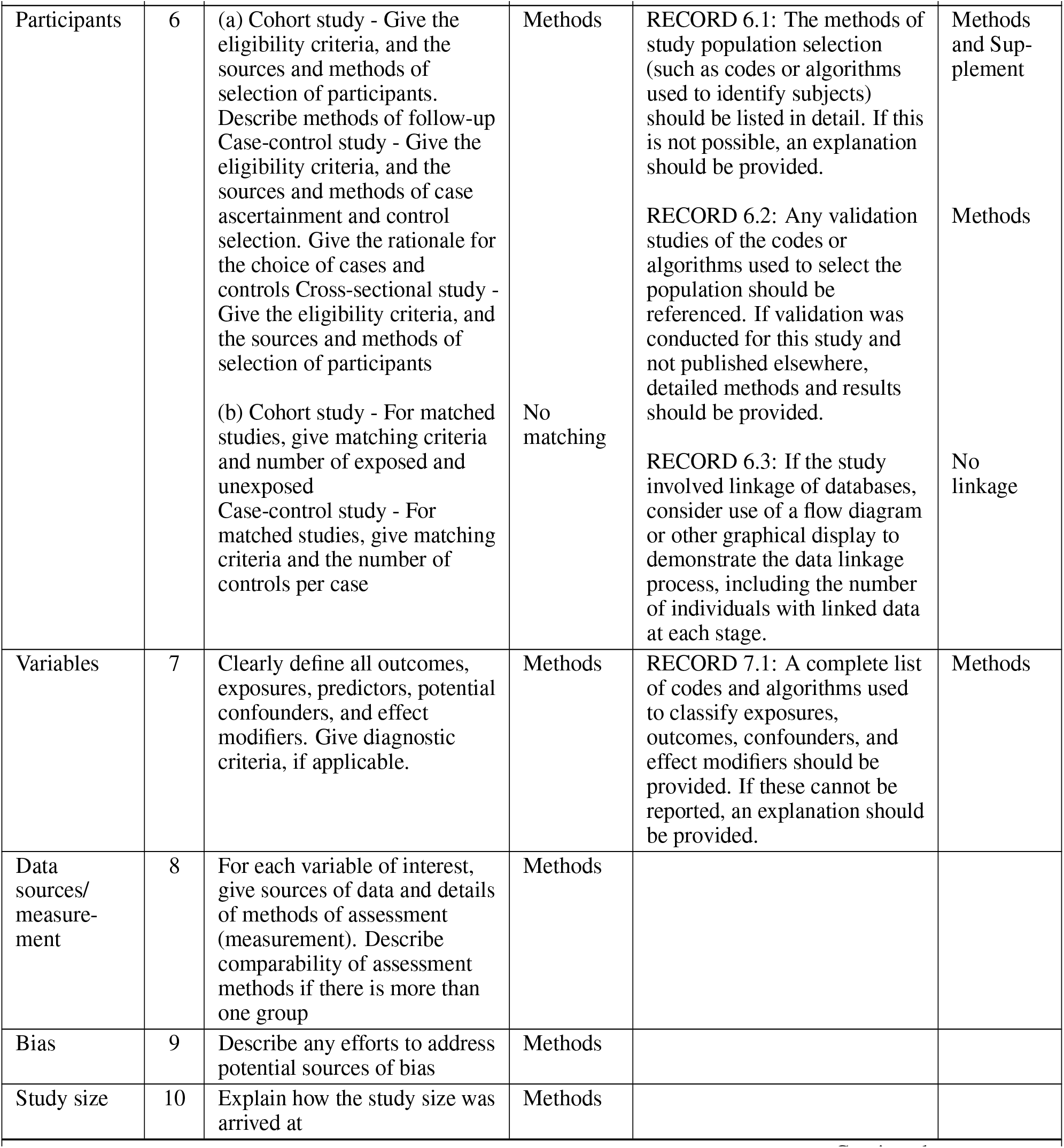
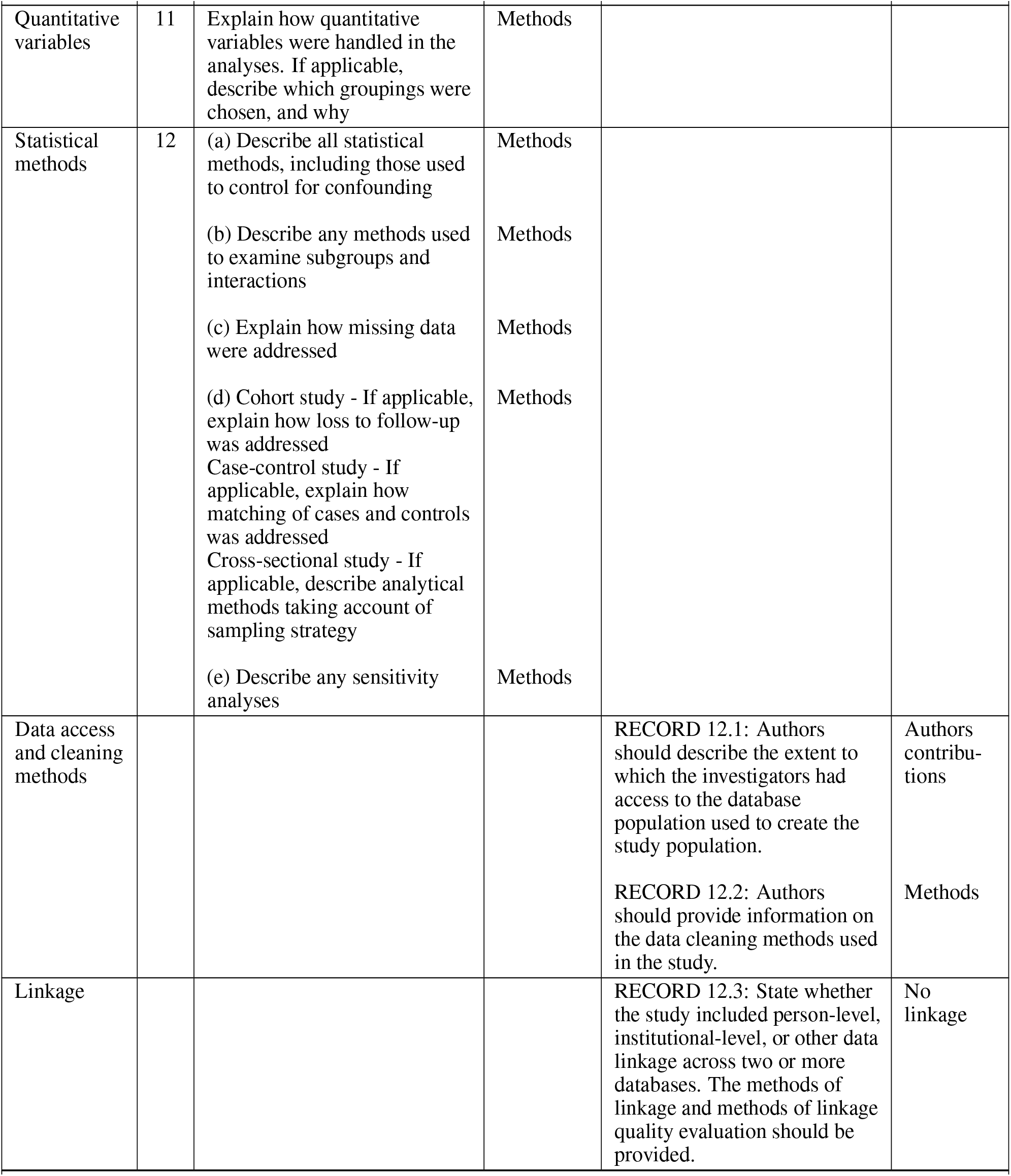
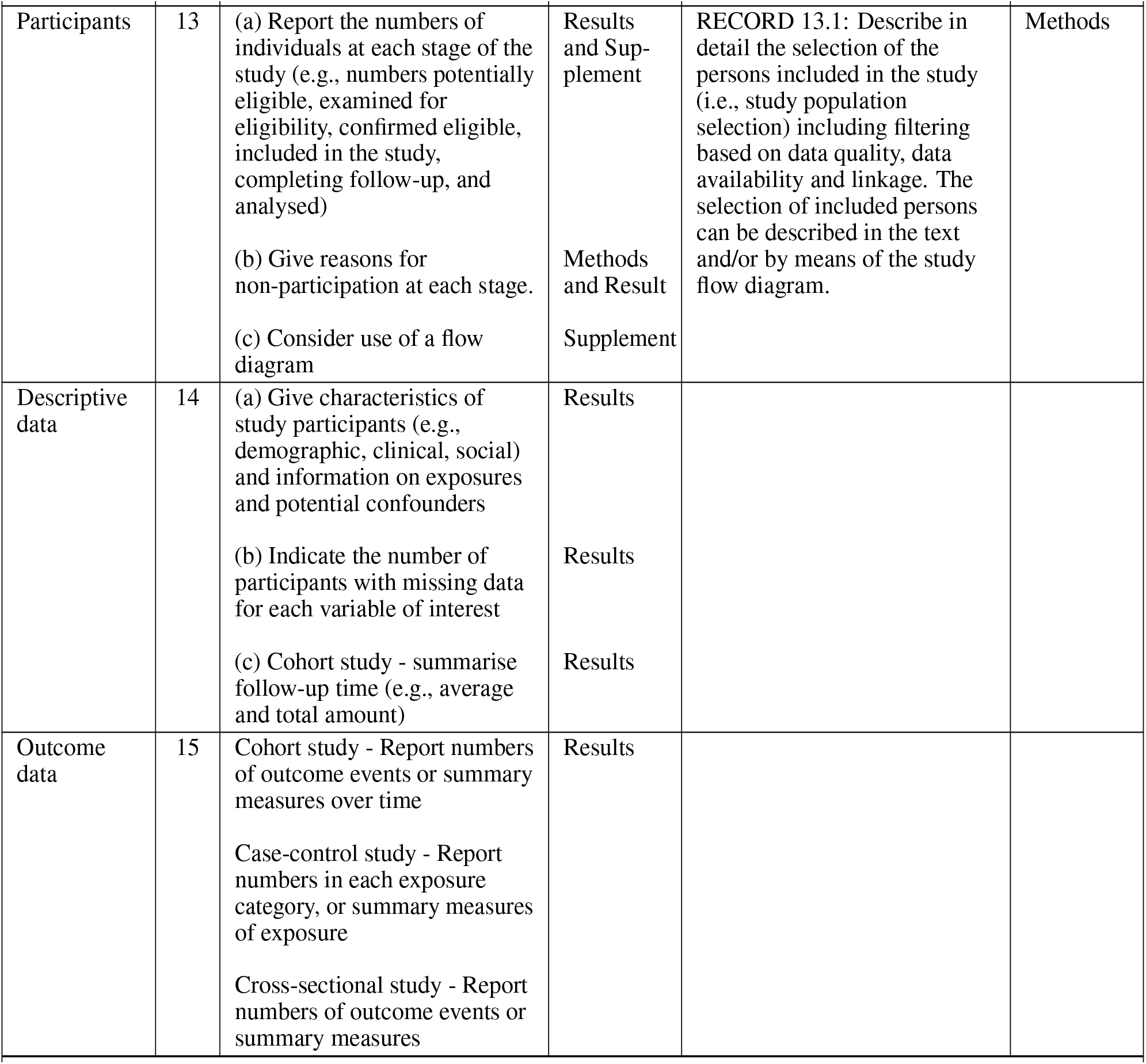
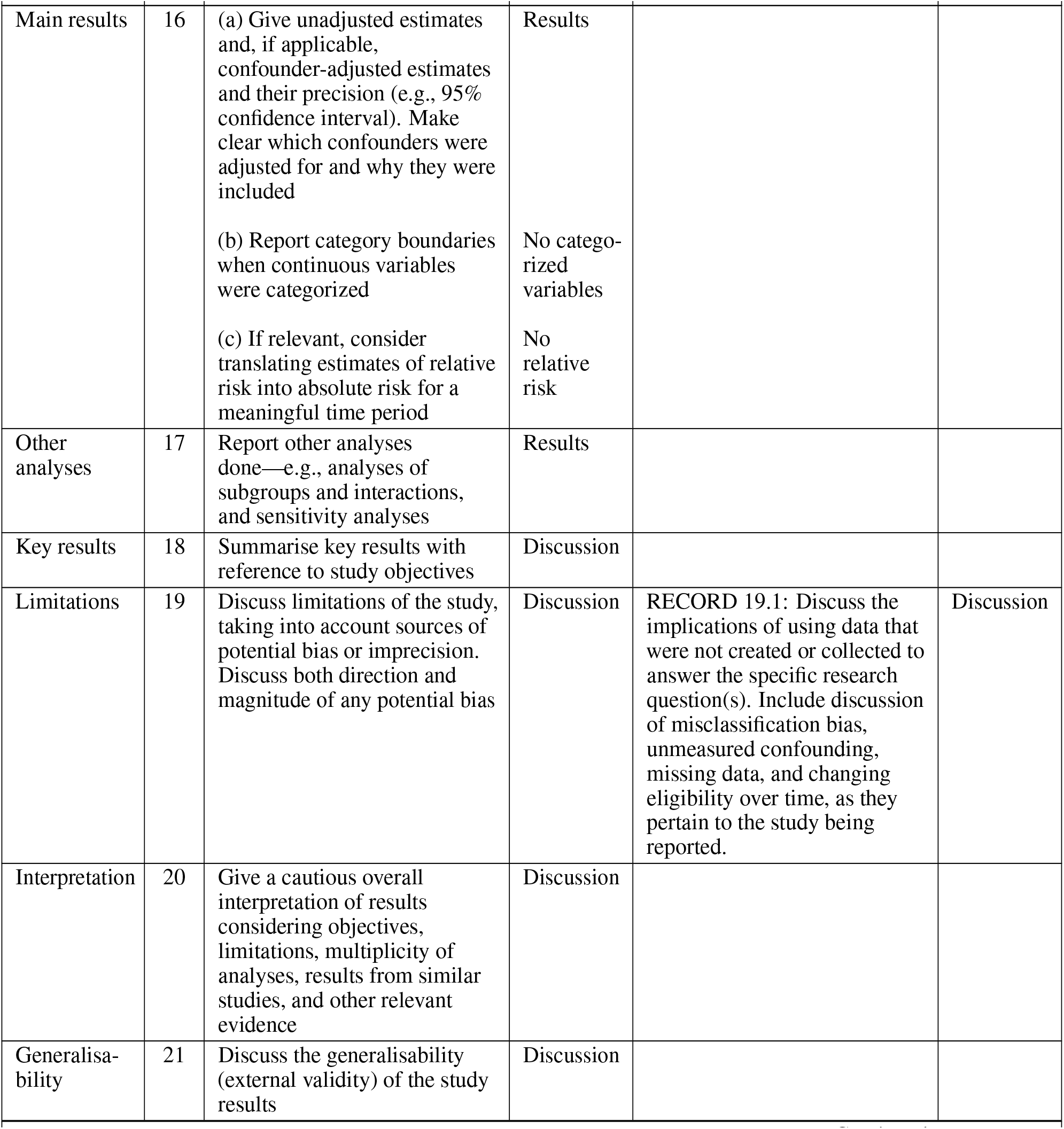
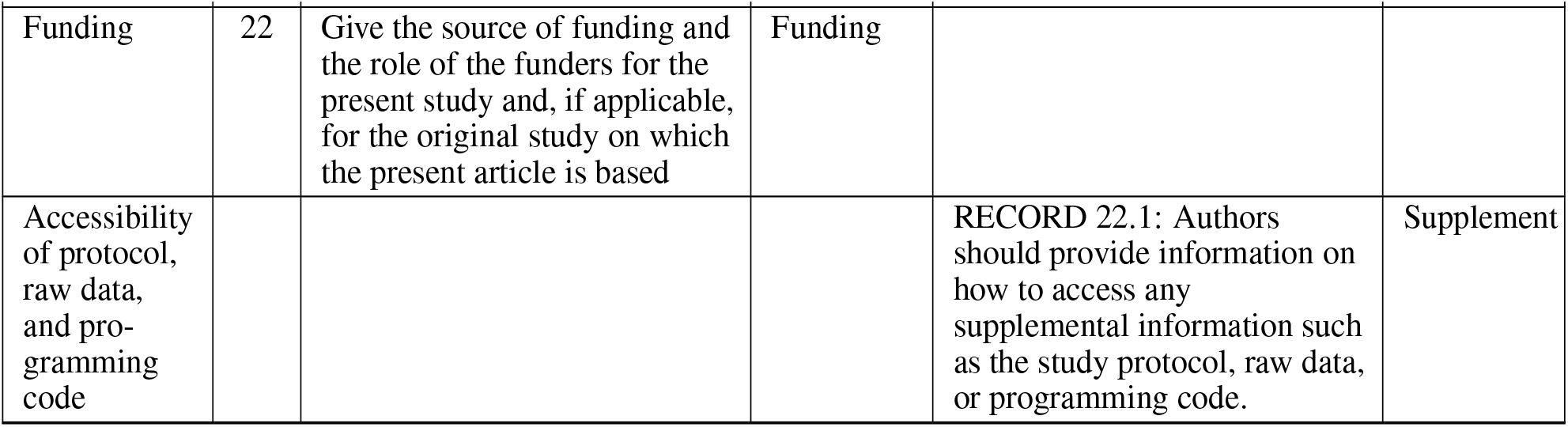

F The RECORD's statement checklist
|  | Item No. | STROBE items | Location in manuscript where items are reported | RECORD items | Location in manuscript where items are reported |
| --- | --- | --- | --- | --- | --- |
| Title and abstract | 1 | (a) Indicate the study's design with a commonly used term in the title or the abstract.<br>(b) Provide in the abstract an informative and balanced summary of what was done and what was found | Abstract<br>Abstract | RECORD 1.1: The type of data used should be specified in the title or abstract. When possible, the name of the databases used should be included.<br>RECORD 1.2: If applicable, the geographic region and timeframe within which the study took place should be reported in the title or abstract.<br>RECORD 1.3: If linkage between databases was conducted for the study, this should be clearly stated in the title or abstract. | Title and abstract<br>Abstract<br>No linkage |
| Background rationale | 2 | Explain the scientific background and rationale for the investigation being reported | Abstract and introduction |  |  |
| Objectives | 3 | State specific objectives, including any prespecified hypotheses | Abstract and introduction |  |  |
| Study Design | 4 | Present key elements of study design early in the paper | Abstract, introduction and methods |  |  |
| Setting | 5 | Describe the setting, locations, and relevant dates, including periods of recruitment, exposure, follow-up, and data collection | Methods |  |  |
| Continued on next page |  |  |  |  |  |
| Participants | 6 | (a) Cohort study - Give the eligibility criteria, and the sources and methods of selection of participants.<br>Describe methods of follow-up<br>Case-control study - Give the eligibility criteria, and the sources and methods of case ascertainment and control selection. Give the rationale for the choice of cases and controls<br>Cross-sectional study - Give the eligibility criteria, and the sources and methods of selection of participants<br>(b) Cohort study - For matched studies, give matching criteria and number of exposed and unexposed<br>Case-control study - For matched studies, give matching criteria and the number of controls per case | Methods<br>No matching | RECORD 6.1: The methods of study population selection (such as codes or algorithms used to identify subjects) should be listed in detail. If this is not possible, an explanation should be provided.<br>RECORD 6.2: Any validation studies of the codes or algorithms used to select the population should be referenced. If validation was conducted for this study and not published elsewhere, detailed methods and results should be provided.<br>RECORD 6.3: If the study involved linkage of databases, consider use of a flow diagram or other graphical display to demonstrate the data linkage process, including the number of individuals with linked data at each stage. | Methods and Supplement<br>Methods<br>No linkage |
| Variables | 7 | Clearly define all outcomes, exposures, predictors, potential confounders, and effect modifiers. Give diagnostic criteria, if applicable. | Methods | RECORD 7.1: A complete list of codes and algorithms used to classify exposures, outcomes, confounders, and effect modifiers should be provided. If these cannot be reported, an explanation should be provided. | Methods |
| Data sources/<br>measurement | 8 | For each variable of interest, give sources of data and details of methods of assessment (measurement). Describe comparability of assessment methods if there is more than one group | Methods |  |  |
| Bias | 9 | Describe any efforts to address potential sources of bias | Methods |  |  |
| Study size | 10 | Explain how the study size was arrived at | Methods |  |  |

**RECORD's statement checklist continued from previous page**
|  | Item No. | STROBE items | Location in manuscript where items are reported | RECORD items | Location in manuscript where items are reported |
| --- | --- | --- | --- | --- | --- |
| Quantitative variables | 11 | Explain how quantitative variables were handled in the analyses. If applicable, describe which groupings were chosen, and why | Methods |  |  |
| Statistical methods | 12 | (a) Describe all statistical methods, including those used to control for confounding<br>(b) Describe any methods used to examine subgroups and interactions<br>(c) Explain how missing data were addressed<br>(d) Cohort study - If applicable, explain how loss to follow-up was addressed<br>Case-control study - If applicable, explain how matching of cases and controls was addressed<br>Cross-sectional study - If applicable, describe analytical methods taking account of sampling strategy<br>(e) Describe any sensitivity analyses | Methods<br>Methods<br>Methods<br>Methods<br>Methods |  |  |
| Data access and cleaning methods |  |  |  | RECORD 12.1: Authors should describe the extent to which the investigators had access to the database population used to create the study population.<br>RECORD 12.2: Authors should provide information on the data cleaning methods used in the study. | Authors contributions<br>Methods |
| Linkage |  |  |  | RECORD 12.3: State whether the study included person-level, institutional-level, or other data linkage across two or more databases. The methods of linkage and methods of linkage quality evaluation should be provided. | No linkage |
| Continued on next page |  |  |  |  |  |
| Participants | 13 | (a) Report the numbers of individuals at each stage of the study (e.g., numbers potentially eligible, examined for eligibility, confirmed eligible, included in the study, completing follow-up, and analysed)<br>(b) Give reasons for non-participation at each stage.<br>(c) Consider use of a flow diagram | Results and Supplement<br>Methods and Result<br>Supplement | RECORD 13.1: Describe in detail the selection of the persons included in the study (i.e., study population selection) including filtering based on data quality, data availability and linkage. The selection of included persons can be described in the text and/or by means of the study flow diagram. | Methods |
| Descriptive data | 14 | (a) Give characteristics of study participants (e.g., demographic, clinical, social) and information on exposures and potential confounders<br>(b) Indicate the number of participants with missing data for each variable of interest<br>(c) Cohort study - summarise follow-up time (e.g., average and total amount) | Results<br>Results<br>Results |  |  |
| Outcome data | 15 | Cohort study - Report numbers of outcome events or summary measures over time<br>Case-control study - Report numbers in each exposure category, or summary measures of exposure<br>Cross-sectional study - Report numbers of outcome events or summary measures | Results |  |  |
| Continued on next page |  |  |  |  |  |
| Main results | 16 | (a) Give unadjusted estimates and, if applicable, confounder-adjusted estimates and their precision (e.g., 95% confidence interval). Make clear which confounders were adjusted for and why they were included<br>(b) Report category boundaries when continuous variables were categorized<br>(c) If relevant, consider translating estimates of relative risk into absolute risk for a meaningful time period | Results<br>No categorized variables<br>No relative risk |  |  |
| Other analyses | 17 | Report other analyses done—e.g., analyses of subgroups and interactions, and sensitivity analyses | Results |  |  |
| Key results | 18 | Summarise key results with reference to study objectives | Discussion |  |  |
| Limitations | 19 | Discuss limitations of the study, taking into account sources of potential bias or imprecision. Discuss both direction and magnitude of any potential bias | Discussion | RECORD 19.1: Discuss the implications of using data that were not created or collected to answer the specific research question(s). Include discussion of misclassification bias, unmeasured confounding, missing data, and changing eligibility over time, as they pertain to the study being reported. | Discussion |
| Interpretation | 20 | Give a cautious overall interpretation of results considering objectives, limitations, multiplicity of analyses, results from similar studies, and other relevant evidence | Discussion |  |  |
| Generalisability | 21 | Discuss the generalisability (external validity) of the study results | Discussion |  |  |
| Continued on next page |  |  |  |  |  |
| Funding | 22 | Give the source of funding and the role of the funders for the present study and, if applicable, for the original study on which the present article is based | Funding |  |  |
| Accessibility of protocol, raw data, and programming code |  |  |  | RECORD 22.1: Authors should provide information on how to access any supplemental information such as the study protocol, raw data, or programming code. | Supplement |

